# The Society of Radiologists in Training (SRT) 2024 Training Survey on the impact of recent changes to training and non-radiologist roles

**DOI:** 10.1101/2024.11.02.24316642

**Authors:** Yakup Kilic, Jordan Colman, Miraen Kiandee, Bhavana Shyamanur, Wee Ping Ngu, Emma Watura

## Abstract

**Introduction:** In 2016, the Royal College of Radiologists (RCR) launched an initiative to address the radiology workforce crisis in the UK, focusing on increasing the number of training places. To address this, the RCR advocated for the introduction of skill mix and expanded imaging training academies. The NHS’s 2023 long-term workforce plan proposed increased integration of medical associate professions (MAPs) in radiology, although this has raised concerns regarding regulation and training quality.

**Methods:** We conducted an online survey targeting UK radiologists to gather insights on their perceptions of training quality, recent modifications to training, and views on non-radiologist roles. The purpose of the study was to evaluate the ways in which these elements affect their training experiences and general contentment.

**Results:** A total of 227 responses were collected, predominantly from residents across all UK training deaneries. 63.9% of trainees expressed satisfaction with their training although this varied, with trainees in new/hybrid academies reporting lower satisfaction. Concerns about training quality were prevalent, attributed to numerous factors such as consultants working from home and increased training numbers. Although opinions on the new curriculum were divided, many commented on the challenges of meeting the new IR competences, and the greater focus on general training was perceived more adversely. Career prospects in radiology were often bleak.

**Conclusion:** The survey results highlighted a multifaceted array of perceptions and concerns amongst radiology residents regarding several factors negatively impacting their training, such as changes to the curriculum and growth of non-radiologist roles.

## 1. Introduction

The Royal College of Radiologists (RCR) set out a decade-long plan in 2016 on the transformation of radiology training. One of the key tenets of the plan included tackling the issue of radiology workforce crisis through building the radiology workforce. The number of radiologists per head of population in the United Kingdom remains much lower than most other European countries.^1^ RCR has successfully lobbied for the increase in Clinical Radiology training numbers in part, through development of radiology training academies.

Workforce issues have previously been tackled in radiology through academies, with Peninsula (South West Imaging Training Academy), Leeds (Leeds and West Yorkshire Radiology Academy) and Norwich (East of England Imaging Training Academy) radiology academies being established in 2005 and later in south Wales (National Imaging Academy Wales) in 2018.^2,3,4,5^ These schemes typically consist of intensive tutorial-based learning for the first 3 years of training (ST1-3). Radiology academies appear to have been a success with them receiving good satisfaction ratings and increasing training capacity, though at a greater financial cost.^6^ Most of the radiology academies have recently changed their name to “Imaging training academies” as they now additionally train non-radiologist workforce groups.^7^

In 2021 many schemes in England, which were traditional apprenticeship models, became part of a new set of radiology academies. There is minimal information available on this in the public domain, other than an online document outlining the Midlands Imaging Training Academy.^8,9^ The authors are aware that this has been implemented in some of their own training programmes, with Kent, Surrey and Sussex (KSS) deanery becoming part of the Southeast imaging academy along with the Wessex deanery, and North-West England deanery becoming part of the North West Imaging Academy. These academies utilise a hybrid model where trainees are on a hospital placement the majority of the time with some online tutorials and access to ultrasound simulation. These new academies are cited to increase training numbers in the region and to be used to train other workforce groups, such as reporting radiographers and sonographers.^9^ According to NHSE (formerly HEE) the regions with academies other than the original three are the; South East Imaging Training Academy, North West Imaging Training Academy, Midlands Imaging Training Academy and London Imaging Training Academy.^7^

Another way proposed to reduce workforce issues by the RCR is by making the radiology training curriculum more general, with 50% of time to be in general radiology during sub-specialty years, and increasing the number of expected interventional competencies.^10^ This is to produce more consultants who can support general and acute radiology.

Skill-mix, a well-known term within radiology, when used appropriately, has proven to be valuable especially in helping provide good patient care. The NHS long term workforce plan, released in 2023, aims to expand extended practices to non-doctor roles in all specialties.^11,12^ This includes a large expansion in newer roles of medical associate professions (MAPs) such as physician associates, which have been introduced into a small number of radiology departments, specifically within interventional radiology. Additionally, the workforce plan cites backlogs in performing and reporting radiology examinations as a key issue. To help reduce this, the expansion of non-doctor roles alongside the increased use of clinical Artificial Intelligence (AI), is thought to be key.

The effect of the expansion of MAP roles has generated significant concerns from several medical organisations, specifically within radiology, we saw the membership of the British Society of Interventional Radiology (BSIR), voting against GMC regulation of MAPs at their 2024 EGM.^13^ The expansion of radiology training numbers and change in curriculum has also been reported to have a negative effect on training. Concerns have been raised recently, mostly echoed on social media, on the definition and scope of MAP’s and their role within the healthcare team to patients and the public. Consequently, we have decided to survey current UK Radiology Trainees/ residents (formerly registrar) on their experience of radiology training overall, recent changes within training and their views on current non-radiologist roles including the proposed expansion.

### Aims

1. Identify trainees’ perception of training quality and assess variations between training schemes.
2. Identify how trainees have been affected by the new RCR curriculum.
3. Identify how trainees have been affected by measures to expand the workforce.
4. Identify how trainees are affected by non-radiologist reporting and interventional roles and their perception of them.
5. Evaluate trainees’ outlook on the future of radiology as a career and future intentions.

## 2. Methods/Materials

Using the online survey tool ‘Google Forms’, a questionnaire was designed to understand the current state of radiology training and how current MAP roles and existing non-doctor roles within radiology are impacting training. After an initial draft of questions were produced, a consensus meeting was held, and the final questionnaire was approved. It included both open and closed ended questions. The questionnaire was open to all radiology trainees, radiology consultants and SAS/non-trainee grade radiology doctors in the UK.

The questionnaire included the following subsections: general training, physician associates, reporting radiographers, sonographers, non-doctors reporting, artificial intelligence and future career prospects within radiology. The questionnaire was distributed via social media platforms including ‘Instagram’ and ‘X’. E-mail to almost all radiology training deaneries were sent out to ask for the questionnaire to be completed by their trainees. All participants were asked if they wanted to answer questions on PA’s, as this was not mandatory to answer the remaining subsections. The questionnaire responses were open from 17^th^ January - 16^th^ February 2024. SRT membership was not mandatory. Data was imported into Excel (Microsoft Excel 2021) for analysis and used to produce all graphs. Significance testing performed using Mann-Whitney U or Spearman’s rank correlation coefficient. Data tables for question responses can be found in the appendix.

No ethical approval was required for this public survey on the experiences of radiologists in training/ radiology residents with national training number (NTN). All respondents provided electronic consent for the storage of their personal data, and for their anonymized responses to be used in future publications and reports.

## 3. Results

### 3.1. Response Demographics and Training Scheme

A total of 227 responses were received. All respondents had declared their GMC numbers. Responses were received from all training deaneries, with the distribution shown in figure 1. The highest number of responses were from London (45). The majority of respondents were trainees, with 25 respondents (11%) being consultants, post-CCT fellows or non-training doctors, their responses were removed from section 3.2 where questions regarding overall training satisfaction.

**Figure 1:**
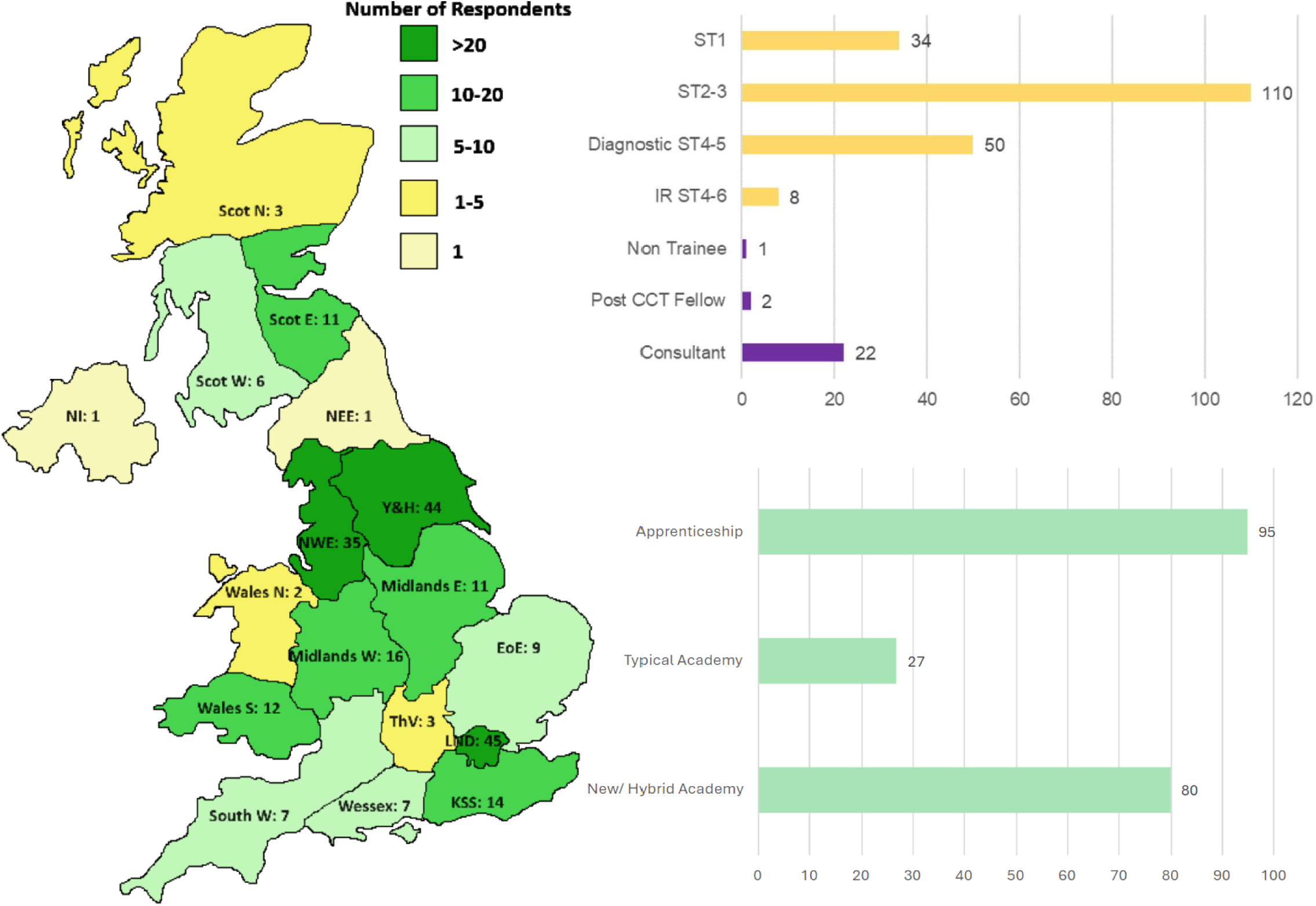
Distribution of responses from all UK deaneries (Left) with number from each deanery shown shaded in colour as per key. The distribution by training grade and non-trainee category is shown in the graph top right, trainees in yellow and non-trainees in purple. Bottom right shows number of responses split by training scheme. Type of training scheme type for each area decided by majority response in that region.

We gathered data from respondents on the type of training scheme they were in (Table 1). Interestingly, all new academies except for North East England (which only had one response) had a large number of conflicting responses with many individuals in these regions reporting they were in a traditional apprenticeship or implied they were unsure in the free text comments.

**Table 1:** Individual training programs with subjective trainee satisfaction score, scheme type and region/academy with regional mean satisfaction score. Ordered by satisfaction score of region. If there were conflicting responses on if the region was an academy the majority response was selected and noted in the 4^th^ column of this.

| Program | Mean Rating | Training scheme Type | Uncertainty if is an academy? | Academy or Region | Mean rating of academy or region |
| --- | --- | --- | --- | --- | --- |
| North East: Newcastle | 5 | New Academy |  | North east imaging training academy | 5 |
| South West: Severn | 4.5 | Apprenticeship |  | South West: Severn | 4.5 |
| Yorkshire and Humber: Leeds | 4.2 | Typical Academy |  | Leeds and west Yorkshire radiology academy | 4.2 |
| Scotland South East: Edinburgh | 4.2 | Apprenticeship |  | Scotland South East: Edinburg | 4.2 |
| London North Central & East: UCL | 4.1 | Apprenticeship |  | London | 4.0 |
| London North Central & East: Royal Free | 5.0 |  |  |  |  |
| London North West: Imperial | 3.7 |  |  |  |  |
| London South: St George's | 4.0 |  |  |  |  |
| London South: GSTT | 3.7 |  |  |  |  |
| London North Central & East: St Bart's & The Royal | 3.5 |  |  |  |  |
| London North West: Northwick Park | 3.5 |  |  |  |  |
| East of England: Norwich | 4.0 | Typical Academy |  | East of England imaging training academy | 4.0 |
| Northern Ireland | 4.0 | Apprenticeship |  | Northern Ireland | 4.0 |
| Wales: North | 4.0 | Apprenticeship |  | Wales: North | 4.0 |
| Yorkshire and Humber: Sheffield | 4.2 | Apprenticeship |  | Yorkshire and Humber | 3.9 |
| Yorkshire and Humber: Hull & East Yorkshire | 3.6 |  |  |  |  |
| Scotland East: Dundee | 3.8 | Apprenticeship |  | Scotland East: Dundee | 3.8 |
| Scotland West: Glasgow | 3.8 | Apprenticeship | Yes | Scotland West: Glasgow | 3.8 |
| Wales: South | 3.7 | Typical Academy |  | National Imaging Training Academy Wales | 3.7 |
| South West: Plymouth & Peninsula | 3.7 | Typical Academy |  | South West Imaging Training Academy | 3.7 |
| Thames Valley: Oxford | 3.7 | Apprenticeship | Yes | Thames Valley | 3.7 |
| Wessex: Southampton | 4.5 | New/ Hybrid Academy | Yes | South East Imaging Training Academy | 3.4 |
| Wessex: Portsmouth | 4.0 |  |  |  |  |
| KSS: Kent | 3.0 |  |  |  |  |
| KSS: Surrey | 3.0 |  |  |  |  |
| KSS: Sussex | 2.5 |  |  |  |  |
| West Midlands: Stoke | 4.0 | New/ Hybrid Academy | Yes | Midlands Imaging Training Academy | 3.3 |
| East Midlands: Nottingham | 3.7 |  |  |  |  |
| West Midlands: Birmingham | 3.1 |  |  |  |  |
| East Midlands: Leicester | 2.3 |  |  |  |  |
| East of England: Essex & Bedfordshire | 3.5 | Apprenticeship |  | East of England | 3.2 |
| East of England: Cambridge | 2.0 |  |  |  |  |
| North West: Lancashire | 3.8 | New/ Hybrid Academy | Yes | North West Imaging Training Academy | 3.1 |
| North West: Manchester | 3.1 |  |  |  |  |
| North West: Mersey | 2.9 |  |  |  |  |
| Scotland North: Aberdeen | 2.3 | Apprenticeship |  | Scotland North: Aberdeen | 2.3 |

### 3.2. General Training Satisfaction and Impact of Recent Training Changes

#### Satisfaction and Training Scheme Type

202 trainee responses were included. Table 1 shows mean scores for satisfaction with training by training program and region/academy. Figure 2 shows most trainees are satisfied with training (63.9%) and further splits satisfaction ratings by the training scheme type and shows regional distributions on a map of the training regions. This demonstrates the new/ hybrid academy having significantly lower satisfaction scores than traditional academies or apprenticeship schemes (P = 0.00052 and P = 0.00008 respectively), with 30.0% being unsatisfied, compared to 6.4% on apprenticeship schemes and 0% in a typical academy. A reasonable number from a hybrid academy, however, are still satisfied with training 45.1%, although lower than the 76.8% from an apprenticeship schemes and 74.1% from a traditional academy.

**Figure 2:**
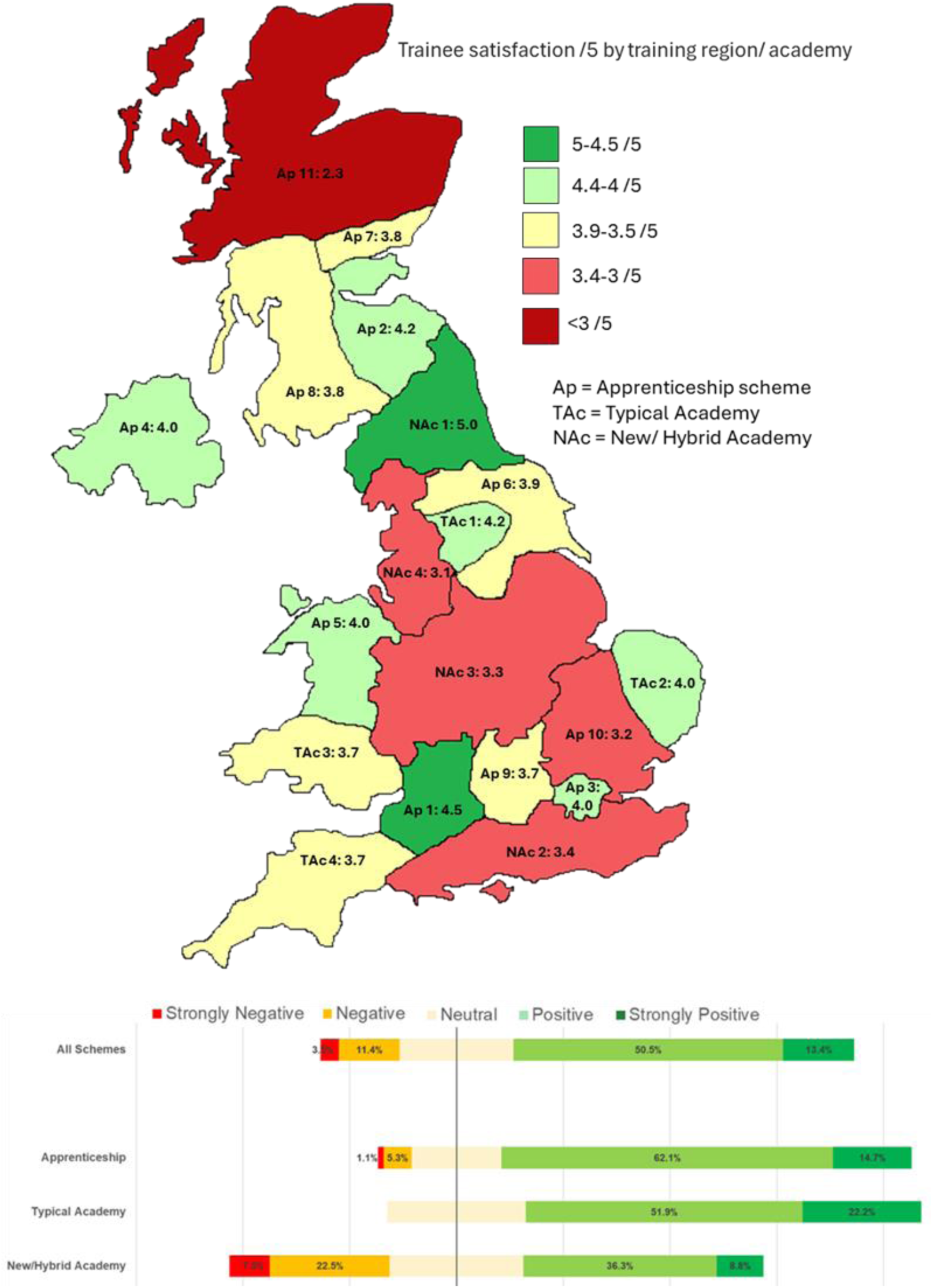
Responses for question” Overall, have you been satisfied with the quality of training in your deanery, so far?” Total responses: 202. Shown for all schemes and split by type of training scheme, shown on a map (top) and as aggregated average bar chart (bottom). Apprenticeship schemes: Ap 1 = South West: Severn, Ap 2= Scotland South East: Edinburg, Ap 3 = London, Ap 4 = Northern Ireland, Ap 5 = Wales: North, Ap 6 = Yorkshire and Humber, Ap 7 = Scotland East: Dundee, Ap 8 = Scotland West: Glasgow, Ap 9 = Thames Valley, Ap 10 = East of England, Ap 11 = Scotland North: Aberdeen. Traditional Academies: TAc 1 = Leeds and west Yorkshire radiology academy, TAc 2 = East of England imaging training academy, TAc 3 = National Imaging Training Academy Wales, TAc 4 = South West Imaging Training Academy. New/ hybrid academies: NAc 1 = North east imaging training academy, NAc 2 = South East Imaging Training Academy, NAc 3 = Midlands Imaging Training Academy, NAc 4 = North West Imaging Training Academy. When compared using a two tailed Mann-Whitney U test the training satisfaction in New/ hybrid academies was significantly lower than traditional academies or apprenticeship schemes (P = 0.00052 and P = 0.00008 respectively). There was no significant difference in satisfaction of traditional academies and apprenticeship schemes (P = 0.589).

While most people are aware on how to raise concerns on training quality (88.1%), only 43.4% of trainees believed that if a concern about training quality was raised, they would be taken seriously (Table 2).

**Table 2.** Table showing responses on raising concerns on training quality.

|  | Yes | No | Unsure |
| --- | --- | --- | --- |
| Do you know how / whom to express concerns about training quality? (202) | 88.1% (178) | 4.5% (9) | 7.4% (15) |
| If you have raised concerns in the past or intend to do so in the future; do you believe that your concerns about training quality will be taken seriously? (182) | 43.4% (79) | 28.0% (51) | 28.6% (52) |

#### New curriculum and other training changes

Figure 3 demonstrates the responses for a range of different general changes that radiology training has been exposed to. The response to the new 2021 RCR radiology curriculum is mixed but slightly more negative overall. For example, 46.6% found the increase in general training in specialty years (ST4/5) to be negative vs 19.8% positive. A change reported to be particularly negative was the increase in consultants working from home (72.3%). The expansion of training numbers was thought to be mostly negative with 44.2% reporting this as negative vs 14.2% positive.

**Figure 3.**
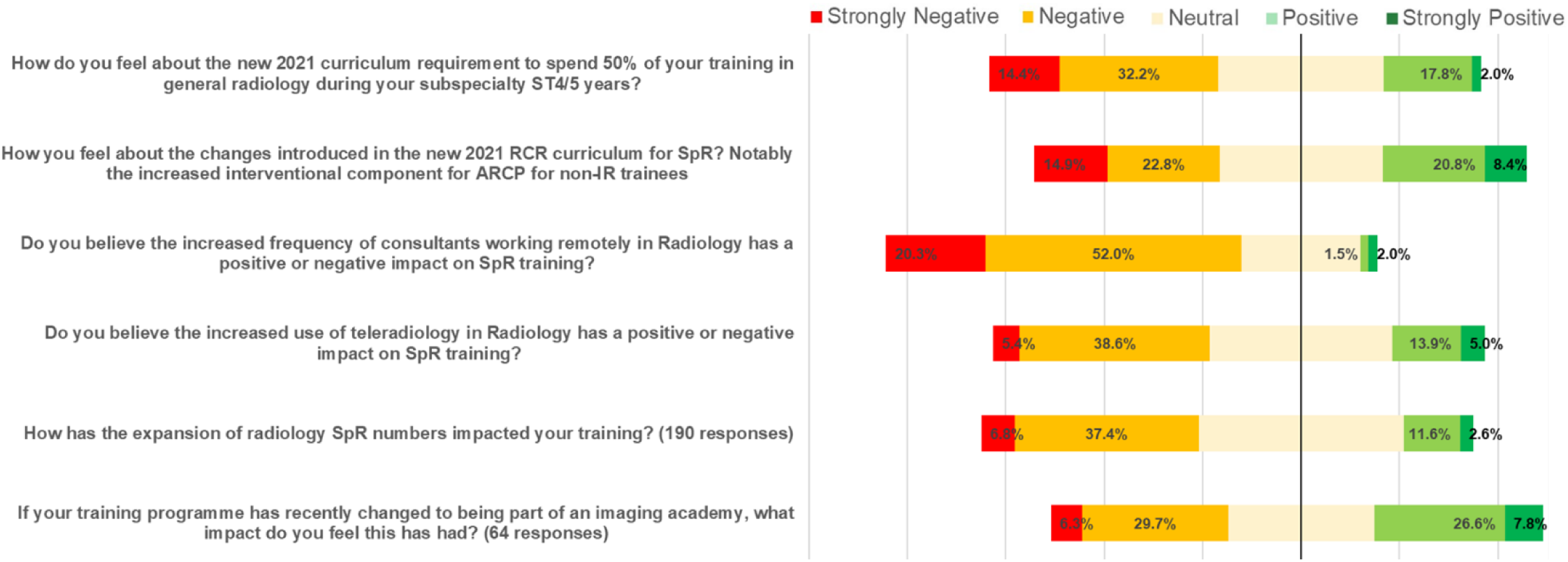
Responses for a range of questions about general changes to training on if they have a negative or positive effect. 202 responses unless otherwise specified with percentages for each response labeled.

Figure 4 shows many respondents agreed that their training region has not adequately prepared for the increase in training numbers (43.1% vs only 16.3% who agree their region had prepared adequately). Many people (40.6%) additionally felt that they would need to do additional fellowship years after training to feel confident in their subspecialty.

**Figure 4.**
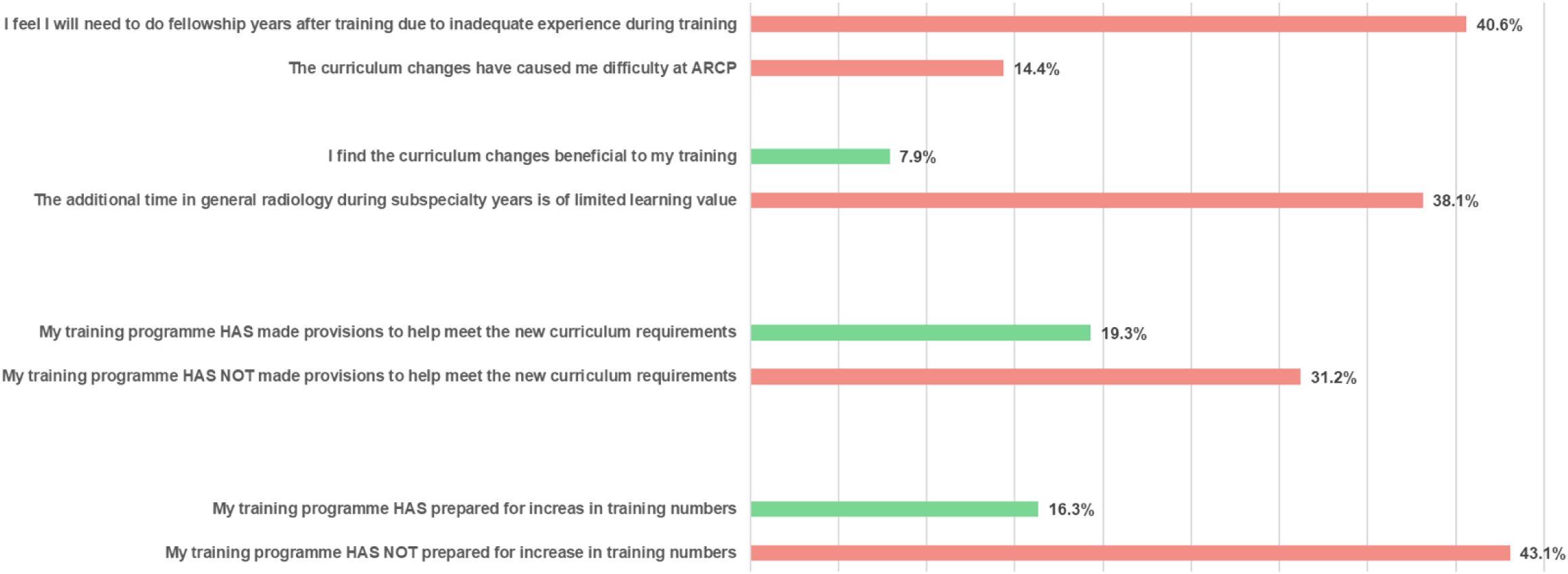
Responses to multiple select questions with list of statements. Positive statements in regards to the topic in green and negative in red.

Respondents were asked, “How often have you been unable to learn to report a study or perform a procedure due to the presence of a non-radiologist role performing or learning that task?”. The responses of which are in figure 5 demonstrate this is happening to the majority of residents at least occasionally and more frequently for many. Figure 6 shows the regional trends. The reported frequency of this happening was found to correlate negatively with training satisfaction (R = -0.213, P = 0.0023) as seen in appendix 2, Figure A2.1.

**Figure 5.**
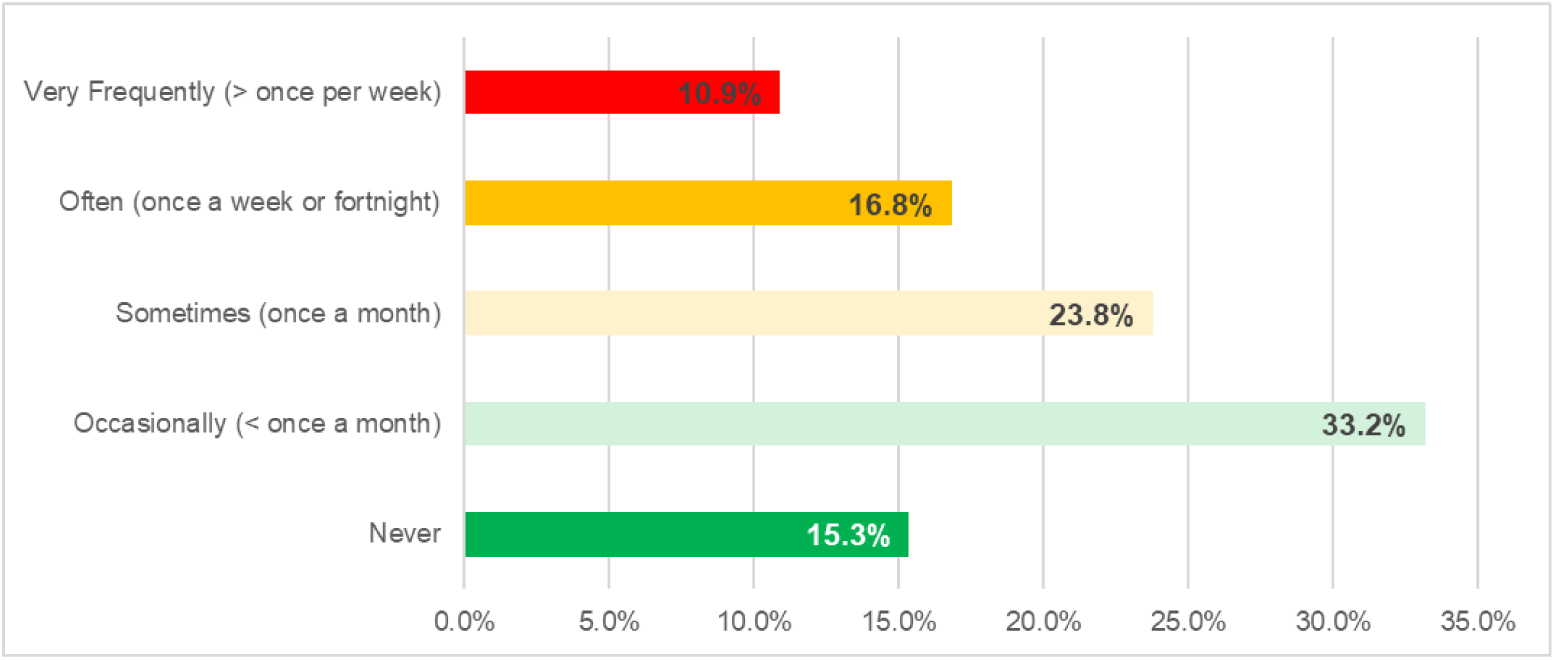
Answers to question “How often have you been unable to learn to report a study or perform a procedure due to the presence of a non-radiologist role performing or learning that task?”. 202 responses.

**Figure 6:**
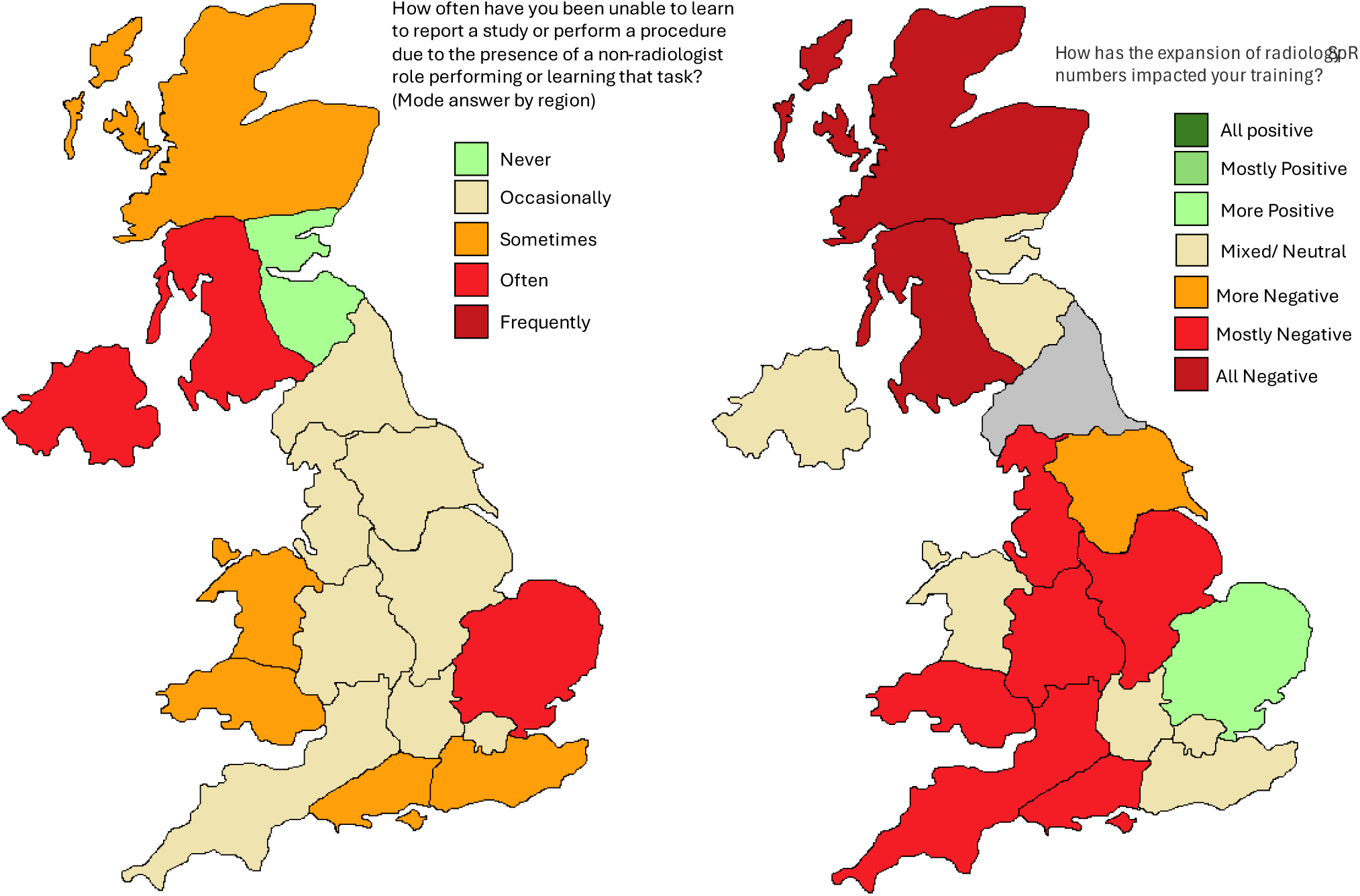
Distribution of responses from all UK deaneries to the above questions. Divided by Regions/ Deaneries are as seen in figure 1. Full data for the responses to these questions can be found in appendix 2. Grey regions do not have data available/ the question is not relevant to region.

In figure 6, regional effects of trainee expansion are shown. How positive the reported impact of the increase in training numbers correlated positively with training satisfaction (R = 0.232, P = 0.0013) as seen in Figure A2.1 (full details in appendix 2).

#### Free Text Comments

Respondents could comment freely on general training issues in free text. Table 4 shows the common issues mentioned. 15 people cited a frequent lack of reporting stations at work, 18 a lack of IR opportunities and a lack of plain films or certain studies was also mentioned frequently.

**Table 3.**
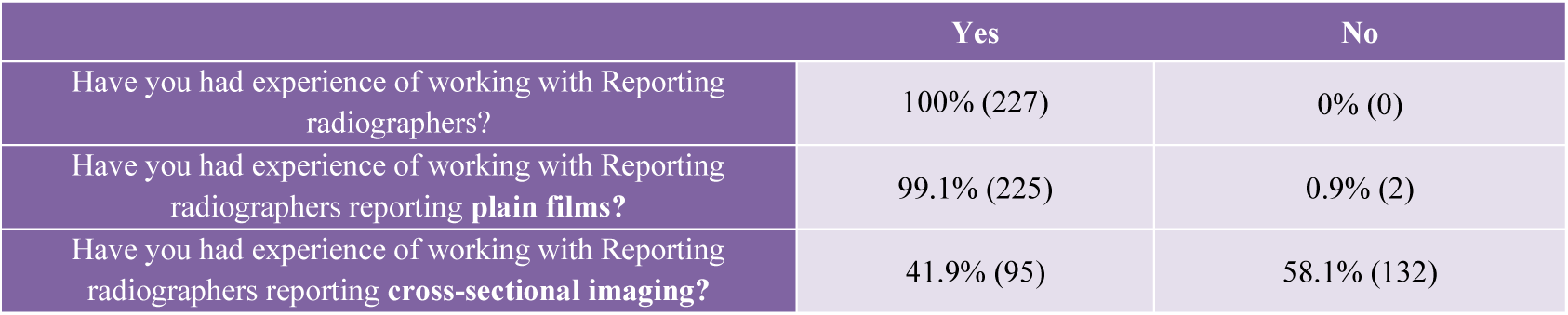
Showing rates of respondents who had worked with reporting radiographers in different capacities.

|  | Yes | No |
| --- | --- | --- |
| Have you had experience of working with Reporting radiographers? | 100% (227) | 0% (0) |
| Have you had experience of working with Reporting radiographers reporting **plain films**? | 99.1% (225) | 0.9% (2) |
| Have you had experience of working with Reporting radiographers reporting **cross-sectional imaging**? | 41.9% (95) | 58.1% (132) |

**Table 4.** Table summarizing free text comment box with commonly mentioned issues listed with number of people who mentioned them. Grouped by issue theme.

| Issues mention in free text comment box | No. who mentioned |
| --- | --- |
| **General training (n=61)** |  |
| Lack of IR opportunities | 18 |
| Lack of reporting stations | 15 |
| lack of certain modality types | 13 |
| lack of plain films making it difficult to reach numbers | 11 |
| lack of consultant time to check reports | 10 |
| Academy teaching being poorly organized or unhelpful | 7 |
| IR or general training requirements in new curriculum taking time from subspecialty training | 4 |
| Being in smaller hospitals with limited training opportunities | 3 |
| **Comments on PAs in radiology (n=79)** |  |
| Concerns regarding patient safety | 12 |
| Unfair when are many Doctors who would like to work in radiology who would be more suited | 15 |
| Inadequate background knowledge to report or preform procedures | 22 |
| Would increase competition for training opportunities | 18 |
| No role in radiology due to minimal ward or administrative work | 10 |
| Inappropriately request scans | 3 |
| No role for PAs as role already filled by sonographers, reporting radiographers and nurses | 18 |
| May be beneficial in some way | 5 |
| **Comments on reporting radiographers (n=88)** |  |
| Creating difficulties in reaching plain film numbers due to lack of studies | 17 |
| Lead to more complex studies or body parts left increasing difficulty reporting | 11 |
| RR should not be able to report cross sectional imaging | 39 |
| Lead to over recommendation of further imaging | 4 |
| Increased competition for studies or learning opportunities | 11 |
| Leads to need for double reporting | 5 |
| Role for RR reducing backlogs | 10 |
| Feel safe plain film reporters | 18 |
| May create more radiographer workforce shortages in non reporting areas | 3 |
| Useful in teaching junior registrars/residents | 7 |
| Unfair when amount of training required is lower | 3 |
| **Sonographers impact on training (n=65)** |  |
| Lack of consultant list impacting training/ training would be better from consultant | 8 |
| Sonographers prioritizing sonography students | 12 |
| Useful teaching from sonographers | 17 |
| Concerns for safety of sonographers performing procedures | 9 |
| **APs impact on training (n=34)** |  |
| Generally good and work inside area of competencies | 4 |
| Helpful and train junior registrars/residents | 5 |
| Concerns regarding lack of medical knowledge to perform procedures | 2 |
| Increased competition for training opportunities | 10 |
| **Non radiologist doctors reporting imaging studies and performing image guided procedures (n=60)** |  |
| Reduces training opportunities as clinicians don't train radiology registrars/residents | 7 |
| Increases work load due to need for double reporting | 3 |
| Uses up radiology resources affecting training e.g. using work stations | 3 |
| Increase competition for learning opportunities | 4 |
| Concerns regarding safety of interpretation of findings outside organ or expertise leading to missed diagnosis or unnecessary further imaging | 9 |
| Concerns on lack of training in medical physics or basic reporting/ imaging principles | 5 |
| **Artificial intelligence impact on training (n=57)** |  |
| Could be/ is a useful aid to improve reporting efficiency or accuracy | 14 |
| Current software of poor quality or accuracy | 24 |
| Can create more work due to call from clinicians reading inaccurate AI finding | 2 |

#### International Medical Graduates (IMGs)

We also asked questions on the specific experience of IMGs entering UK training schemes. The full details are in appendix 3. Briefly, 46 respondents report they were IMGs with the majority (60.0%) reporting positively on support for integration.

### 3.3. Impact on Non-radiologist Roles in Radiology on Training

#### Physicians Associates (PAs)

Of the 227 responses 24 reported that they had experience of working with PAs while working in radiology. If the respondents had not worked with PAs while in radiology, they were asked to report what they thought the effect would be. The responses were mostly negative regardless of whether the respondents had worked with PAs or not as shown in Figure 7. The strongly negative or negative response rate for their suspected effect on training being 95.9% if they had experience with working with PAs and 96.3% if they hadn’t. Figure 8 additionally shows statements the respondents agree with, with a small number agreeing with positive responses such as “PAs should be trained to interpret and report plain films” (8) vs large numbers which agree with negative responses such as “There is no role for PAs interpreting or reporting diagnostic imaging or performing image guided procedures” (144).

**Figure 7:**
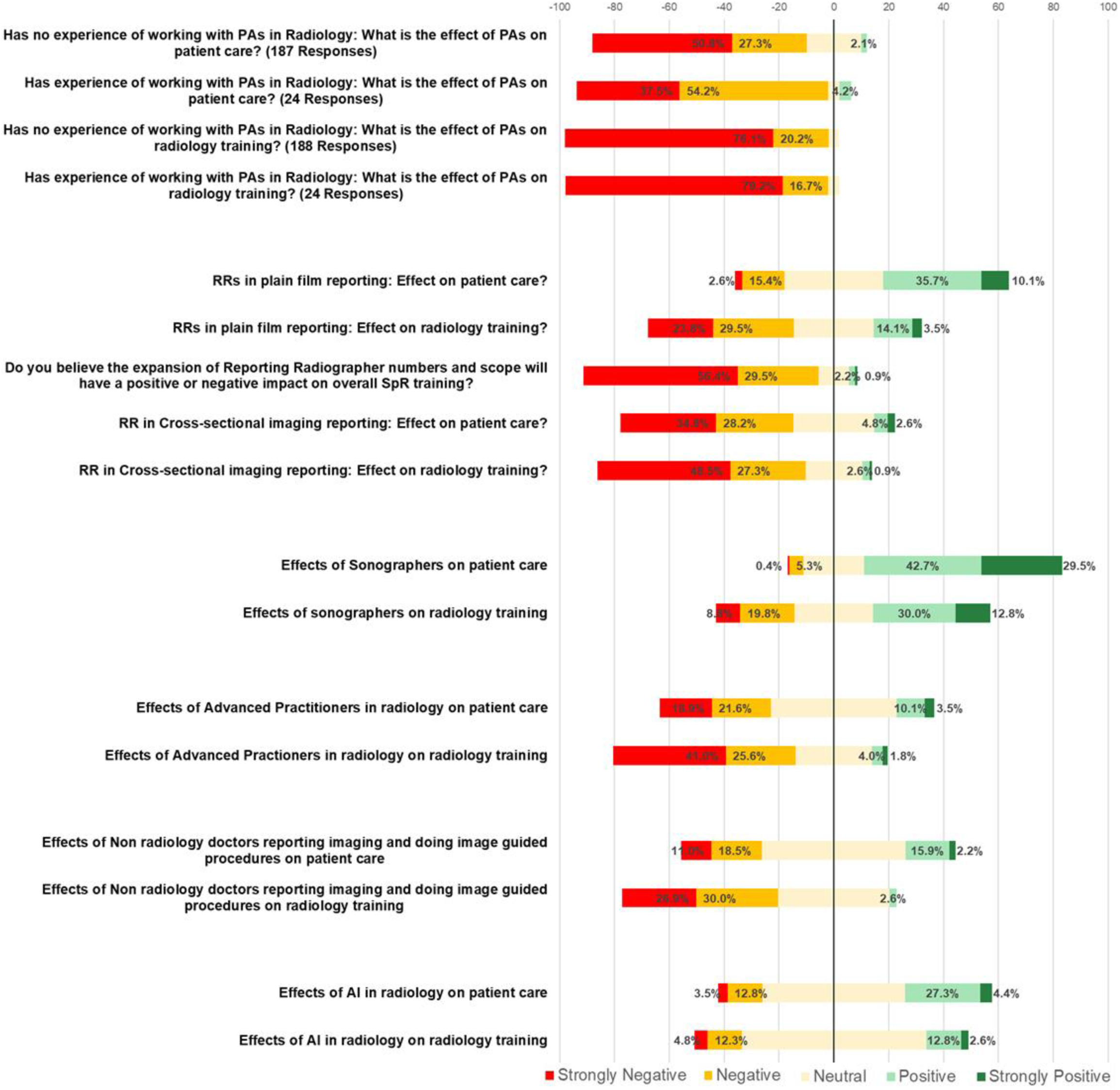
Responses on how different non-radiologist roles reporting and undertaking advanced practice affect training with percentages shown for number of strongly negative (red), negative (orange), positive (light green) or strongly positive (dark green) responses. They are grouped by role.

**Figure 8.**
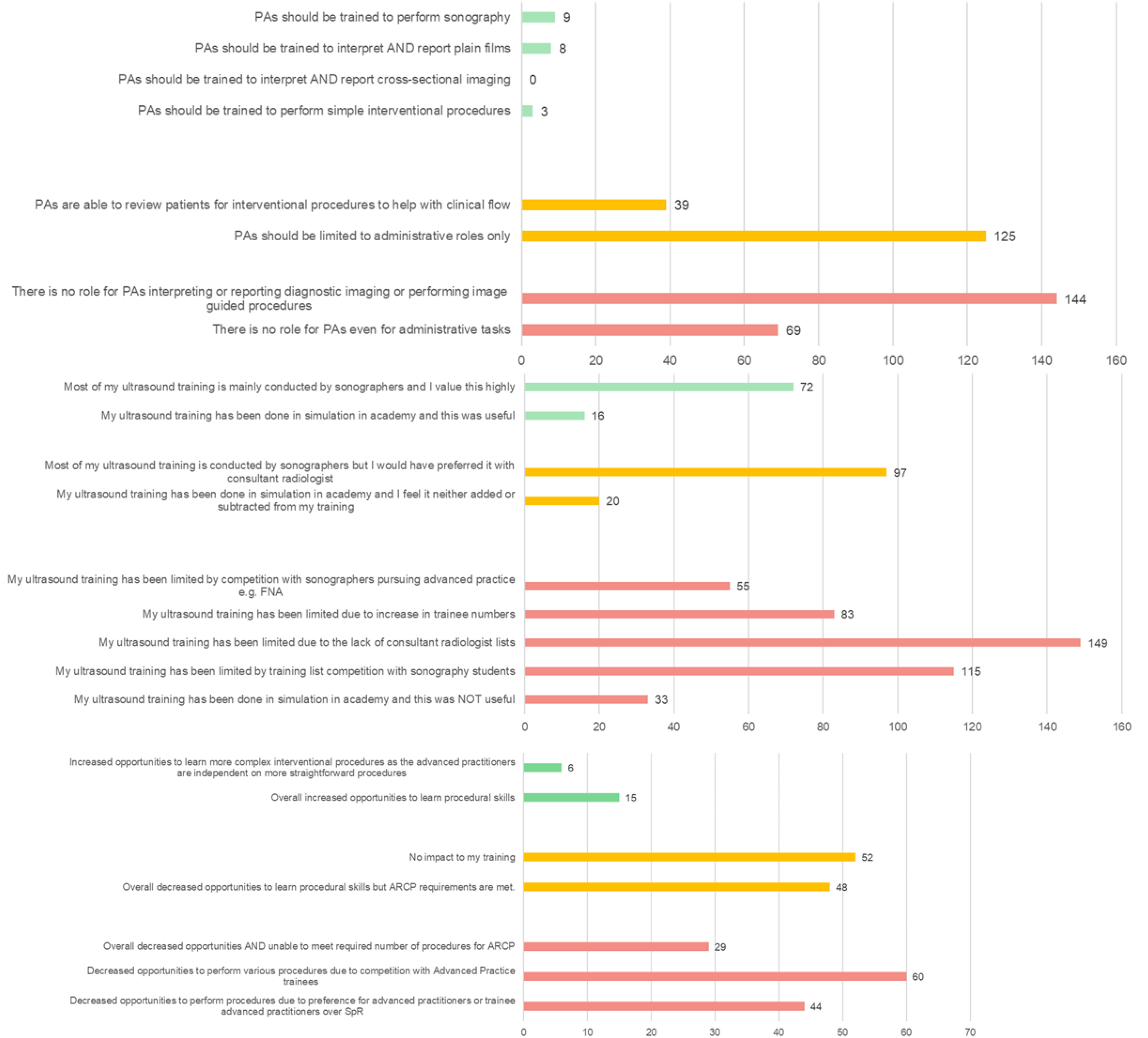
Responses to multiple select questions with list of statements on non-radiologists roles on radiology training. Positive statements on the topic are in green, neutral yellow and negative red. Top bar chart is regarding statements pertaining to Pas, the middle bar chat to statements regarding sonographers or ultrasound in general and the bottom bar chart is in regards to advance practitioners.

Furthermore, respondents were able to give free text comments on the topic with a summary of common topics mentioned in table 4. We report 22 people mentioning concerns about the inadequate background knowledge to report imaging studies or perform procedures, 10 people raising the fact that there is inadequate ward or administrative work for them to do in radiology and 18 people mentioning that radiographers, sonographers or nurses already fill these roles.

#### Reporting Radiographers

As seen in table 3 all respondents have worked with reporting radiographers in some form. 41.9% of which reported this being in cross-sectional imaging.

Most respondents felt reporting radiographers have a positive impact on patient care in reporting plain films with 45.8% reporting a positive effect (Figure 7). The responses to the impact on radiology training, however, are more mixed with 53.3% reporting a negative effect and 17.6% positive. There is a mostly negative response to the effect of their expansion on training (85.9% negative). Cross sectional imaging reporting by radiographers were particularly viewed negatively. As seen in the comments in table 4 this may be due to concerns radiographers not having enough medical background knowledge (mentioned by 39 people). Lastly, there are comments effects on training with competition for studies reported to be causing issues with reaching required plain film numbers (17) and broadly there is an increased competition for select studies and other opportunities (11).

#### Sonographers

Only 3 respondents had not worked with sonographers. Figure 7 shows a mostly positive effect of sonographers on patient care (72.2% positive or very positive) although, it is more split on their effect on training. 28.6% report a negative effect vs 42.8% positive. In figure 8, 72 people agree that they value training from sonographers highly, although many people agreed that training had been limited due to competition with sonographer students (115). This may be exacerbated by increase in Radiology trainee numbers (statement agreed with by 83) and lack of consultant list (149). In the free text comments summarised in table 4, while many commented on beneficial training from sonographers (17) it is also commented some people have sonography students prioritised over them (mentioned by 12) and concerns are raised that sonographers do not have adequate background medical knowledge to perform procedures (9).

#### Advanced Practitioners in IR

97 of respondents (42.7%) reported having worked with advanced nurse practitioners or radiographer practitioners within interventional radiology. Figure 7 reports their effect on training and patient care with the responses being mostly negative. We found that many people agree that the presence of these staff members lead to decreased opportunities to learn procedures (60), although 52 agree with the statement they have no impact on training and 15 agree with the statement that they increase opportunities to learn procedural skills (see figure 8).

#### Non-Radiologist Doctors

126 respondents (55.5%) had experience of working with non-radiologist doctors reporting imaging or performing image guided procedures. The impact on training and patient care is displayed in figure 7 with 63.4% of respondents reported concerns in the expansion of this role.

#### AI

137 respondents (60.4%) reported some experience with AI in radiology. The impact of this shown in figure 7 with it being very mixed. In table 4 there are many comments mentioning that current software is of poor quality or accuracy (24 people mentioned).

### 3.4. Future Outlook

207 respondents (91.2%) answered questions on the outlook of radiology with responses shown in figure 9. Overall, the reported outlook was poor with 61.2% reporting a pessimistic outlook on the future of radiology as a career. Non-radiologist roles expanding in radiology appeared to play a role with 77.8% reporting their expansion to make them more likely to leave the UK and 55.1% reporting they would not be comfortable at all in supervising non-radiologists report or perform procedures and 82.0% reporting any degree of discomfort with this.

**Figure 9.**
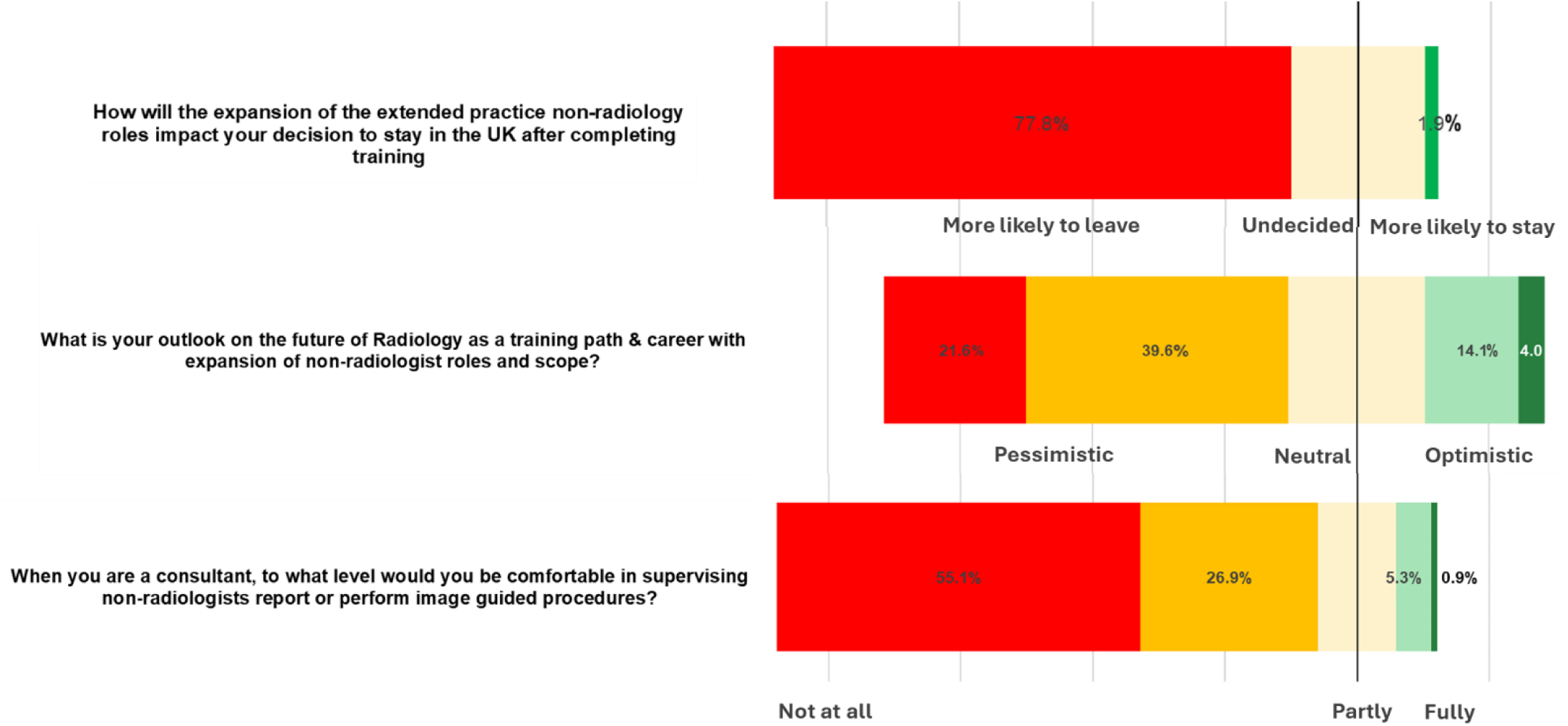
Response for questions on future outlook.

## 4. Discussion

### General Training

Overall, there remains a high satisfaction rate with radiology training. However, we have also shown a markedly reduced satisfaction with training schemes which have recently become new/ hybrid academies. The new academies appear to have been created due to the success of the original academies, however, their high training quality and satisfaction scores are not replicated. An explanation could be that the teaching is typically online which has been found to be rated lower in quality compared to face-to-face teaching in some studies.^14,15^ In contrast, however, some individuals (34.4%) did report a positive impact of the transfer to an academy, although this was less than those who reported a negative effect of the transfer (36%).

We showed several recent changes reported as having a negative impact on training, in particular the increased frequency of consultants working from home and expansion of training numbers overall appears to have a negative effect on training. Lack of workstations, plain films to report or other specific studies were additionally frequently cited as issues.

The effects of the new curriculum are mixed with it being split on if the new interventional requirements are positive. More people reported the increased general training requirement during sub-specialty years to be negative, likely as most feel it to be of limited learning value. Many additionally feel they will need to do further post-training fellowship time to be confident in their subspecialty, which may delay many individuals in becoming consultants and mitigate any reductions in workforce shortages from the new curriculum, especially in specialist areas. A large number of individuals did report that their training programme had not adequately prepared for the curriculum changes with people commenting that meeting the new IR requirements were difficult.

### Non-Radiologist roles

We surveyed views on numerous non-doctor roles reporting and performing procedures in radiology and their expansion. Some of these more established roles such as plain film reporting radiographers and sonographers were mostly viewed as positive for patient care. However, their expansion of scope such as reporting radiographers reporting cross-sectional imaging and sonographers performing procedures was view mostly negatively, likely due to concerns raised about the lack of appropriate medical training for these more complex tasks. Additionally, the effect of these roles on training was mixed, with increased competition for learning opportunities likely being a large factor.

Less well-established roles such as PAs and advanced practitioners are viewed mostly negatively both for the effect on patient care and radiologist training. Only a small number of respondents had experience of working with PAs in radiology and many of the responses likely speculative, but even when subdivided to only those who have experience, the responses remain almost entirely negative. It is additionally raised that PAs likely lack the medical knowledge to report or perform procedures safely and that other roles in the radiology department are already adequately filled more appropriately.

The role of non-radiologist doctors in radiology was also of concern being viewed mostly negatively. This may be due to concerns of potential dilution of specialized training as despite their medical background, may not have the same depth of training in imaging principles and interpretation.

### Future outlook

The perception of radiology as a career in the future overall was very pessimistic. There was a reported marked unwillingness to supervise non-radiologist roles and the expansion of these roles meaning individuals are much more likely to leave after training. As outlined in many comments, this is likely due to the increased difficulty of training and supervising roles with less background knowledge. It may be because this is expected to require more time and supervision and the supervisor is likely to incur increased medicolegal risk. This suggests that the plan to reduce workload by introducing and expanding these non-radiologist roles will likely be negated by an exodus of trained radiologist and subsequently a lack of qualified Radiologist to supervise them.

### 4.1. Limitations

While our survey aimed to capture a comprehensive overview of the experiences and opinions of current UK Radiology Trainees regarding their training and the proposed expansion of non-radiologist roles, several limitations must be acknowledged. The survey was distributed to current UK Radiology Trainees, but the response rate was low and the total number of respondents were not uniform across all regions and training levels. This may result in a sample that is not fully representative of the entire population of radiology trainees in the UK. Participants who chose to respond to the survey may have had stronger opinions or specific experiences related to the training changes and the expansion of MAP roles. This self-selection bias could skew the results toward more extreme viewpoints and may not accurately reflect the broader trainee population’s perspectives. The survey was conducted at a particular time when the expansion of MAP roles and changes in the curriculum were highly topical. This temporal context may have influenced the respondents’ views, which could differ if the survey were conducted at a different time.

### 4.2. Recommendations

- Training schemes should consider ways of mitigating training difficulties due to consultants working from home and make proper investments and plans for the increased number of trainees.
- Training schemes should review the current process for trainees to meet their interventional competencies and work to improve these.
- A review of the need for such a large proportion of subspecialty years to be spent on general radiology and outline ways to improve its learning value.
- A review of the effectiveness of new imaging training academies and why these are rated more negatively.
- The RCR and training programs to ensure radiology resident training is prioritised and not compromised by the time taken to train non-radiologist roles.
- Key stakeholders to review and strategize how radiology training post can be expanded while maintaining high training quality.

## 5. Conclusion

In summary, the survey results highlight a complex landscape of perceptions and concerns among radiology trainees in the UK regarding various factors affecting their training, including curriculum changes, the expansion of non-radiologist roles, and the outlook of the field. Overall radiology training has a high satisfaction rate, although there is clearly a perceived issue with some areas transferring into new academies.

## Data Availability

All data produced in the present work are contained in the manuscript
All data produced in the present study are available upon reasonable request to the authors

## Appendix

## A.1. General training data tables

**Table A1.1.** Reponses to question: How has the expansion of radiology SpR numbers impacted your training? (total 202 trainee responses).

|  |  |
| --- | --- |
| Very positive | 5 |
| Positive/ some positive | 22 |
| No impact/ neutral | 79 |
| Negative/ some negative | 71 |
| Very negative | 13 |
| N/A | 12 |

**Table A1.2.** Reponses to question: Do you believe the increased use of teleradiology in Radiology has a positive or negative impact on SpR training? (1 = very negative; 5 =very positive) (total 202 trainee responses).

|  |  |
| --- | --- |
| Very positive 5 | 10 |
| Positive/ some positive | 28 |
| No impact/ neutral | 75 |
| negative/ some negative | 78 |
| Very negative 1 | 11 |

**Table A1.1.** Reponses to question: Do you believe the increased frequency of consultants working remotely in Radiology has a positive or negative impact on SpR training? (1 = very negative; 5 =very positive) (total 202 trainee responses)

|  |  |
| --- | --- |
| Very positive 5 | 4 |
| positive/ some positive | 3 |
| No impact/ neutral | 49 |
| negative/ some negative | 105 |
| Very negative 1 | 41 |

**Table A1.1.**
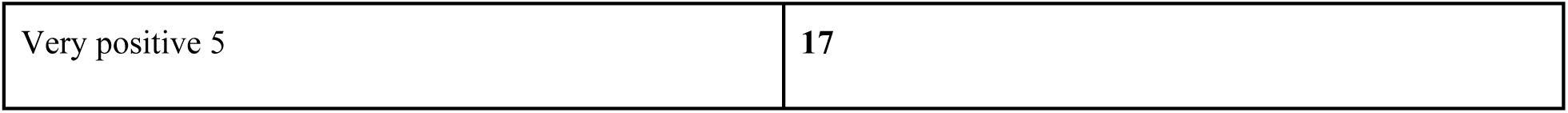
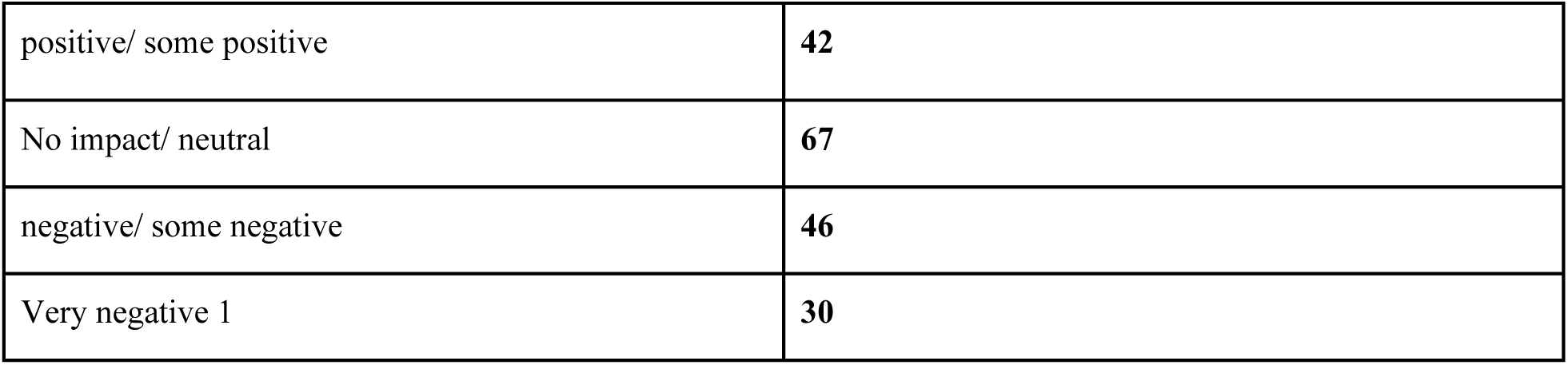
Reponses to question: How you feel about the changes introduced in the new 2021 RCR curriculum for SpRNotably the increased interventional component for ARCP for non-IR trainees (1 = very negative; 5 =very positive) (total 202 trainee responses)

|  |  |
| --- | --- |
| Very positive 5 | 17 |
| positive/ some positive | **42** |
| No impact/ neutral | **67** |
| negative/ some negative | **46** |
| Very negative 1 | **30** |

**Table A1.1.** Reponses to question: How do you feel about the new 2021 curriculum requirement to spend 50% of your training in general radiology during your subspecialty ST4/5 years? (1 = very negative; 5 =very positive) (total 202 trainee responses)

|  |  |
| --- | --- |
| Very positive 5 | **4** |
| positive/ some positive | **36** |
| No impact/ neutral | **68** |
| negative/ some negative | **65** |
| Very negative 1 | **29** |

## A.2. Trainee Satisfaction by region and correlations

**Figure A2.1.**
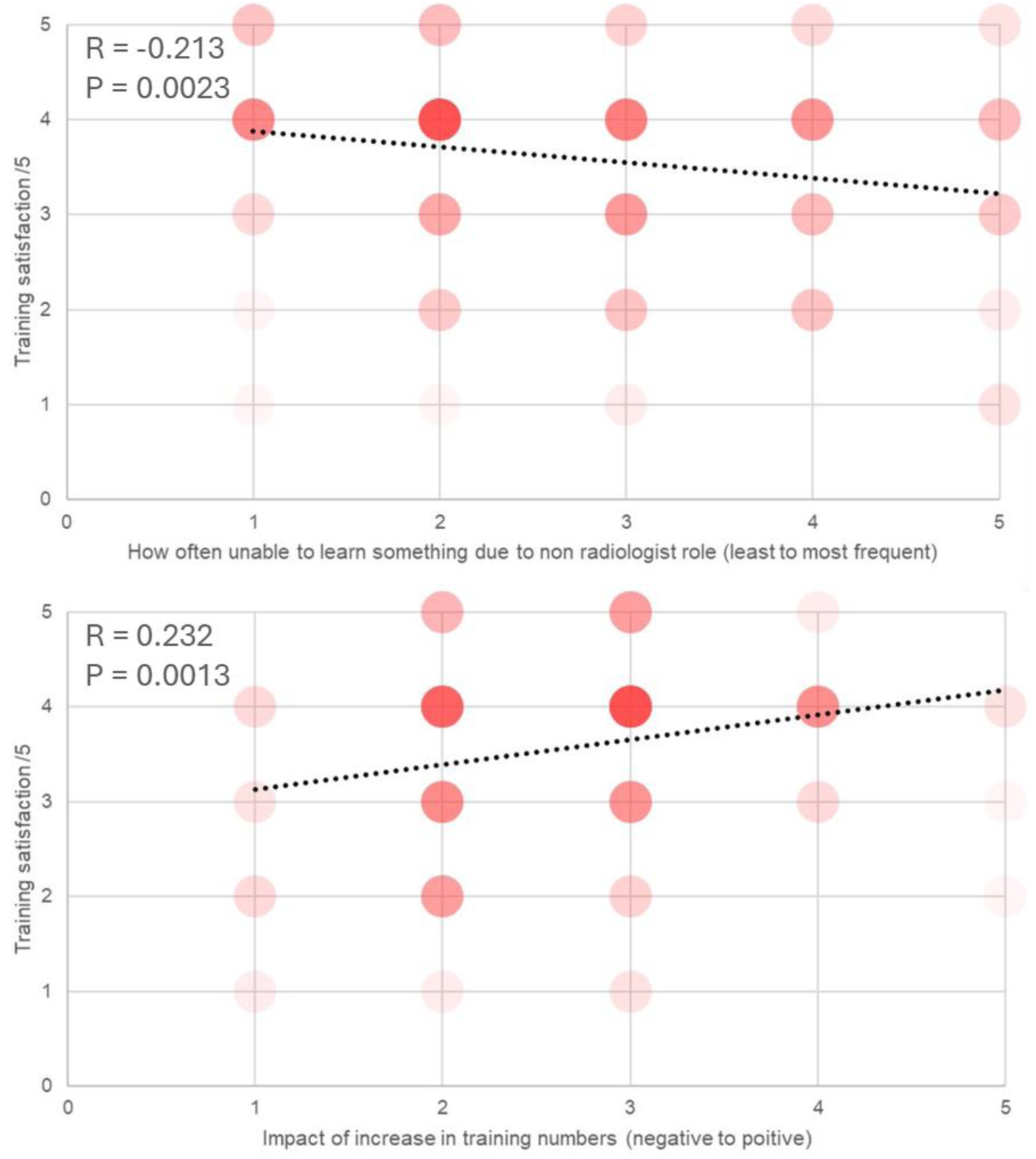
Scatter plot with number of data points at each value displayed by density. Plot of Training satisfaction with Frequency of being unable to learn to report a study or perform a procedure due to the presence of a non-radiologist role performing or learning that task (top) and with the impact of increased training numbers on training (bottom) with trend line, R and significance of correlation shown. For training satisfaction 1 = Very negative, 2 = Negative, 3 = neutral, 4 = positive and 5 = Very positive. For frequency of being unable to learn to report a study or perform a procedure due to the presence of a non-radiologist role performing or learning that task 1 = Never, 2 = occasionally (< once a month), 3 = sometimes (once a month), 4 = often (once a week or fortnight_ and 5 = frequently (> once a week). For impact of increase in training numbers to training 1 = Very negative, 2 = Negative, 3 = neutral, 4 = positive and 5 = Very positive. Correlation and significance calculated using spearman’s rank correlation coefficient.

**Table A2.1.** Number of responses to the above question by training program.

|  | How often have you been unable to learn to report a study or perform a procedure due to the presence of a non-radiologist role performing or learning that task? |  |  |  |  |
| --- | --- | --- | --- | --- | --- |
| Training Program | Very Frequently (> once per week) | Often (once a week or fortnight) | Sometimes (once a month) | Occasionally (< once a month) | Never |
| East Midlands: Leicester | 1 | 0 | 1 | 0 | 0 |
| East Midlands: Nottingham | 1 | 1 | 1 | 3 | 1 |
| East of England: Cambridge | 0 | 0 | 1 | 0 | 0 |
| East of England:<br>Essex &<br>Bedfordshire | 0 | 2 | 0 | 1 | 1 |
| East of England:<br>Norwich | 0 | 1 | 1 | 0 | 1 |
| KSS: Kent | 0 | 0 | 0 | 0 | 1 |
| KSS: Surrey | 1 | 0 | 5 | 4 | 0 |
| KSS: Sussex | 0 | 0 | 1 | 1 | 0 |
| London North<br>Central & East:<br>UCL | 4 | 1 | 6 | 9 | 4 |
| London North<br>Central & East:<br>Royal Free | 0 | 0 | 0 | 1 | 0 |
| London North<br>West: Imperial | 0 | 4 | 1 | 0 | 1 |
| London South: St<br>George's | 0 | 1 | 1 | 0 | 0 |
| London South:<br>GSTT | 0 | 0 | 1 | 1 | 0 |
| London North<br>Central & East: St<br>Bart's & The Royal | 1 | 1 | 0 | 1 | 1 |
| London North<br>West: Northwick<br>Park | 0 | 0 | 1 | 0 | 1 |
| North East:<br>Newcastle | 0 | 0 | 0 | 1 | 0 |
| North West:<br>Manchester | 2 | 2 | 4 | 5 | 0 |
| North West:<br>Mersey | 2 | 6 | 1 | 8 | 1 |
| North West:<br>Lancashire | 0 | 2 | 0 | 2 | 0 |
| Northern Ireland | 0 | 1 | 0 | 0 | 0 |
| Scotland East:<br>Dundee | 0 | 0 | 0 | 0 | 5 |
| Scotland North:<br>Aberdeen | 0 | 1 | 1 | 0 | 1 |
| Scotland South<br>East: Edinburgh | 0 | 0 | 0 | 5 | 1 |
| Scotland West:<br>Glasgow | 1 | 2 | 1 | 1 | 1 |
| South West:<br>Plymouth &<br>Peninsula | 0 | 0 | 0 | 2 | 0 |
| South West:<br>Severn | 0 | 0 | 1 | 0 | 1 |
| Thames Valley:<br>Oxford | 0 | 0 | 1 | 1 | 1 |
| Wales: North | 0 | 1 | 1 | 0 | 0 |
| Wales: South | 2 | 1 | 5 | 2 | 1 |
| Wessex:<br>Southampton | 1 | 0 | 3 | 1 | 1 |
| Wessex:<br>Portsmouth | 0 | 0 | 0 | 1 | 0 |
| West Midlands:<br>Birmingham | 1 | 2 | 2 | 5 | 1 |
| West Midlands:<br>Stoke | 1 | 1 | 0 | 2 | 0 |
| Yorkshire and<br>Humber: Hull &<br>East Yorkshire | 2 | 1 | 2 | 4 | 0 |
| Yorkshire and<br>Humber: Leeds | 1 | 2 | 2 | 4 | 2 |
| Yorkshire and<br>Humber:<br>Sheffield | 1 | 1 | 4 | 2 | 3 |

**Table A2.2.** Number of responses to the above question by training program.

|  | How has the expansion of radiology SpR numbers impacted your training? |  |  |  |  |
| --- | --- | --- | --- | --- | --- |
| Program | Very negative impact | Negative / Some negative impact | No impact / Neutral | Positive / Some positive impact | Very positive impact |
| East Midlands:<br>Leicester | 1 | 0 | 0 | 0 | 0 |
| East Midlands:<br>Nottingham | 1 | 4 | 2 | 0 | 0 |
| East of England:<br>Cambridge | 0 | 1 | 0 | 0 | 0 |
| East of England:<br>Essex &<br>Bedfordshire | 0 | 1 | 1 | 2 | 0 |
| East of England:<br>Norwich | 0 | 0 | 2 | 1 | 0 |
| KSS: Kent | 0 | 1 | 0 | 0 | 0 |
| KSS: Surrey | 0 | 3 | 2 | 3 | 1 |
| KSS: Sussex | 0 | 0 | 2 | 0 | 0 |
| London North<br>Central & East:<br>UCL | 0 | 5 | 15 | 1 | 0 |
| London North<br>Central & East:<br>Royal Free | 0 | 0 | 0 | 1 | 0 |
| London North<br>West: Imperial | 0 | 1 | 3 | 2 | 0 |
| London South: St<br>George's | 0 | 0 | 1 | 0 | 0 |
| London South:<br>GSTT | 0 | 1 | 1 | 0 | 0 |
| London North<br>Central & East: St<br>Bart's & The Royal | 0 | 0 | 2 | 1 | 1 |
| London North<br>West: Northwick<br>Park | 0 | 0 | 2 | 0 | 0 |
| North East:<br>Newcastle | 0 | 0 | 0 | 0 | 0 |
| North West:<br>Manchester | 1 | 9 | 3 | 0 | 0 |
| North West:<br>Mersey | 4 | 5 | 6 | 1 | 1 |
| North West:<br>Lancashire | 0 | 3 | 1 | 0 | 0 |
| Northern Ireland | 0 | 0 | 1 | 0 | 0 |
| Scotland East:<br>Dundee | 0 | 1 | 3 | 0 | 0 |
| Scotland North:<br>Aberdeen | 1 | 2 | 0 | 0 | 0 |
| Scotland South<br>East: Edinburgh | 0 | 1 | 1 | 4 | 0 |
| Scotland West:<br>Glasgow | 2 | 4 | 0 | 0 | 0 |
| South West:<br>Plymouth &<br>Peninsula | 0 | 1 | 1 | 0 | 0 |
| South West:<br>Severn | 0 | 1 | 1 | 0 | 0 |
| Thames Valley:<br>Oxford | 0 | 1 | 1 | 1 | 0 |
| Wales: North | 0 | 0 | 1 | 0 | 0 |
| Wales: South | 0 | 9 | 1 | 0 | 0 |
| Wessex:<br>Southampton | 0 | 4 | 2 | 0 | 0 |
| Wessex:<br>Portsmouth | 0 | 0 | 1 | 0 | 0 |
| West Midlands:<br>Birmingham | 0 | 5 | 5 | 0 | 1 |
| West Midlands:<br>Stoke | 1 | 1 | 2 | 0 | 0 |
| Yorkshire and<br>Humber: Hull &<br>East Yorkshire | 0 | 1 | 4 | 3 | 1 |
| Yorkshire and<br>Humber: Leeds | 2 | 3 | 5 | 1 | 0 |
| Yorkshire and<br>Humber:<br>Sheffield | 0 | 3 | 7 | 0 | 0 |

## A.3. International Medical graduates

The question “Are you an international medical graduate (IMG)?” was asked and results showing in table 3.

**Table A3.1.** Responses when asking if respondents were international medical graduates (IMGs).

| Answer to questions “Are you an international graduate (IMG)?” | Number |
| --- | --- |
| Yes and came to UK for radiology training | 3 |
| Yes and worked as a doctor in the UK prior to radiology training | 43 |
| No | 181 |

20% (46/228) of trainees who responded were international medical graduates. The responses for support in integration are shown in figure 7 and was reported to be mostly positive.

**Figure A3.1.**
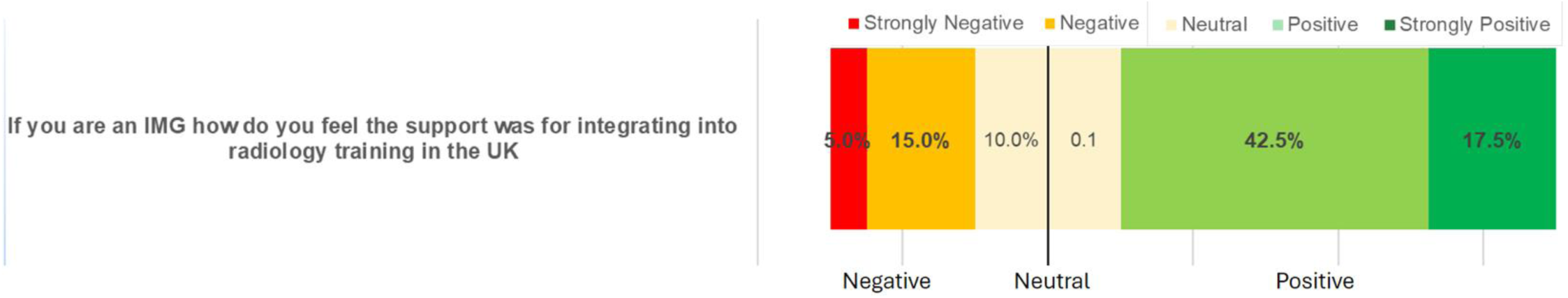
Of the 46 IMGs 40 responded to the question on support for integrating into the UK displayed here.

**Table A3.2.** Reponses to question: If you are an IMG how do you feel the support was for integrating into radiology training in the UK (optional) (1 = poor, 5= excellent)

|  |  |
| --- | --- |
| **1 = Poor** | 2 |
| **2** | 6 |
| **3** | 8 |
| **4** | 17 |
| **5 = Excellent** | 7 |

**Table A3.3.** Table summarizing free text comment box on experience of IMG’s within radiology training

| Free text comments on experience as an IMG | Number of people who mentioned |
| --- | --- |
| Schemes rarely know what an IMG would be. Not prepared for IMGs at all. No special provisions for IMGs. | 1 |
| Didn't feel any difference. Had done medical training in UK prior to radiology and radiology training is much better. But still there are a lot of deficiencies. | 1 |
| I have worked in Canada and training was more supported as training opportunity were not taken up by non-medical health care professionals | 1 |
| No issues as an IMG. Probably because I am white and English speaking. | 1 |
| I'd like to note that my experiences as an IMG will probably differ due to being an EEA graduate. I feel well supported and do not feel that being an IMG has had any negative impacts on my training. | 1 |

## A.4. Impact Non Radiologist Roles data tables

**Table A4.1.**
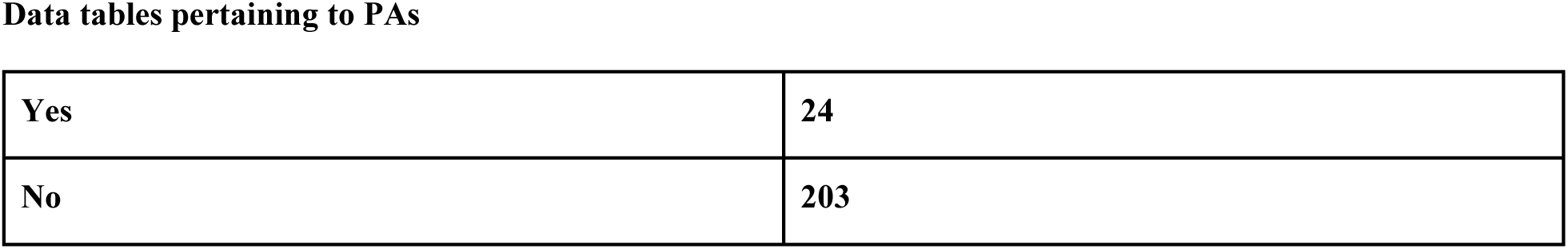
Reponses to question: Have you had experience of working with Physician Associates in Radiology?

**Table A4.2.** Reponses to question: Would you like to answer further questions about the expansion of PA in Radiology ( Diagnostic and Intervention).

|  |  |
| --- | --- |
| **Yes** | **211** |
| **No** | **16** |

**Table A4.3.** Reponses to question: Effect of PAs on Patient care? (1=very negative; 5=very positive) 211 responses

|  |  |
| --- | --- |
| **1 = Very negative** | **104** |
| **2** | **64** |
| **3** | **38** |
| **4** | **5** |
| **5 = Very positive** | **0** |

**Table A4.4.** Reponses to question from those who have not worked with PAs: Effect of PAs on SpR Training? (1=very negative; 5=very positive) 211 responses.

|  |  |
| --- | --- |
| **1 = Very negative** | **162** |
| **2** | **41** |
| **3** | **8** |
| **4** | **0** |
| **5 = Very positive** | **0** |

**Table A4.5.** Reponses to question Of those that answered yes to experience with PA in radiology: Effect on PAs on patient care (24 responses).

|  |  |
| --- | --- |
| **1 = Very negative** | **9** |
| **2** | **13** |
| **3** | **1** |
| **4** | **1** |
| **5 = Very positive** | **0** |

**Table A4.6.** Reponses to question of those that answered yes to experience with PA in radiology: Effect on PAs on SpR training (24 responses).

|  |  |
| --- | --- |
| **1 = Very negative** | **16** |
| **2** | **3** |
| **3** | **1** |
| **4** | **0** |
| **5 = Very positive** | **0** |

Data tables pertaining to reporting radiographers.

**Table A4.7.** Reponses to question: Effect of reporting radiographers reporting plain film imaging on patient care? (1=very negative; 5=very positive) Total respondents 226

|  |  |
| --- | --- |
| **1 = Very negative** | **5** |
| **2** | **35** |
| **3** | **82** |
| **4** | **81** |
| **5 = Very positive** | **23** |

**Table A4.8.** Reponses to question: Effect of reporting radiographers reporting plain films on SpR Training? (1=very negative; 5=very positive)

|  |  |
| --- | --- |
| **1 = Very negative** | **53** |
| **2** | **67** |
| **3** | **66** |
| **4** | **32** |
| **5 = Very positive** | **8** |

**Table A4.9.** Reponses to question: Effect of reporting radiographers reporting cross sectional imaging on patient care? (1=very negative; 5=very positive)

|  |  |
| --- | --- |
| **1 = Very negative** | **79** |
| **2** | **64** |
| **3** | **67** |
| **4** | **11** |
| **5 = Very positive** | **6** |

**Table A4.10.** Reponses to question: Effect of Reporting radiographers reporting cross sectional imaging on SpR Training? (1=very negative; 5=very positive)

|  |  |
| --- | --- |
| **1 = Very negative** | **110** |
| **2** | **62** |
| **3** | **47** |
| **4** | **6** |
| **5 = Very positive** | **2** |

**Table A4.11.** Reponses to question: Do you believe the expansion of Reporting Radiographer numbers and scope will have a positive or negative impact on overall SpR training?

|  |  |
| --- | --- |
| **1 = Very negative** | **129-1** |
| **2** | **67** |
| **3** | **25** |
| **4** | **5** |
| **5 = Very positive** | **2** |

Data tables for questions pertaining to sonographers.

**Table A4.12.** Reponses to question: Have you had experience working with sonographers?

|  |  |
| --- | --- |
| **Yes** | **224** |
| **No** | **3** |

**Table A4.13.** Reponses to question: Effect of sonographers on patient care? (1=very negative; 5=very positive)

|  |  |
| --- | --- |
| **1 = Very negative** | **1** |
| **2** | **12** |
| **3** | **50** |
| **4** | **97** |
| **5 = Very positive** | **67** |

**Table A4.14.** Reponses to question: Effect of sonographers on SpR Training?(1=very negative; 5=very positive)

|  |  |
| --- | --- |
| **1 = Very negative** | **20** |
| **2** | **45** |
| **3** | **65** |
| **4** | **68** |
| **5 = Very positive** | **29** |

**Table A4.15.**
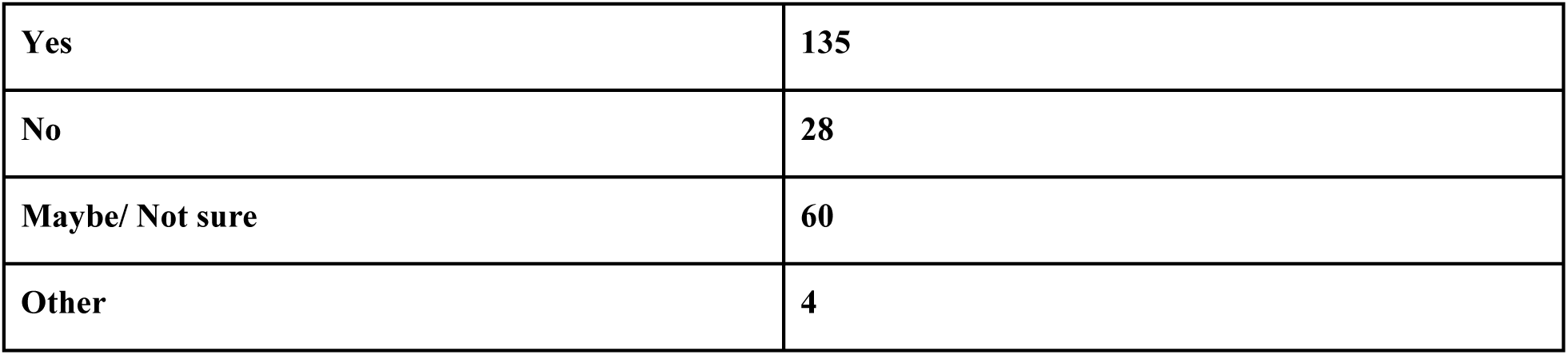
Reponses to question: Are you concerned about the potential impact on training by the continued expansion of sonographer roles including e.g. performing intervention?

|  |  |
| --- | --- |
| **Yes** | **135** |
| **No** | **28** |
| **Maybe/ Not sure** | **60** |
| **Other** | **4** |

Data tables of questions pertaining to Advance practitioners in IR.

**Table A4.16.**
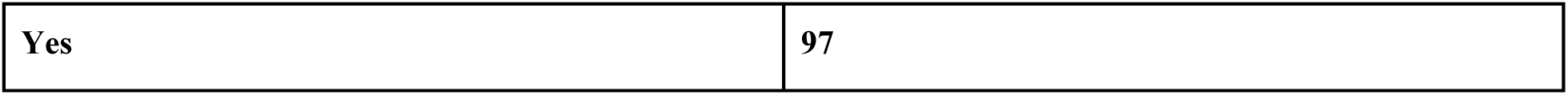
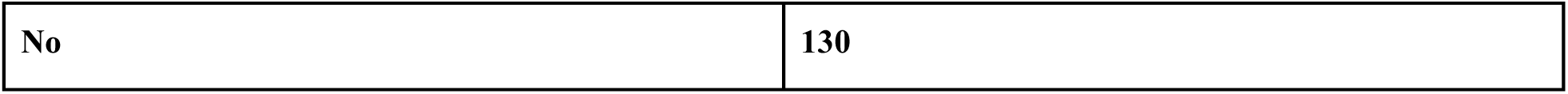
Reponses to question: Have you had experience with Advanced Practitioners in Radiology/IR/INR?

**Table A4.17.**
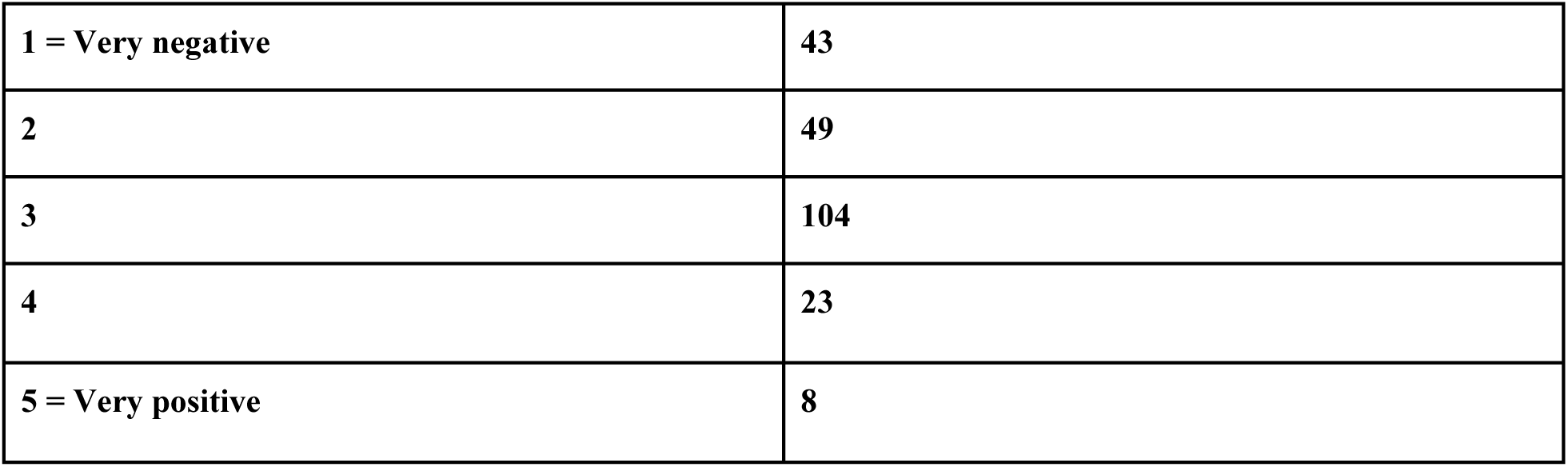
Reponses to question: Effect of advance practitioners on patient care? (1=very negative; 5=very positive)

|  |  |
| --- | --- |
| **1 = Very negative** | **43** |
| **2** | **49** |
| **3** | **104** |
| **4** | **23** |
| **5 = Very positive** | **8** |

**Table A4.17.** Reponses to question: Effect of advance practitioners SpR Training? (1=very negative; 5=very positive)

|  |  |
| --- | --- |
| **1 = Very negative** | **93** |
| **2** | **58** |
| **3** | **63** |
| **4** | **9** |
| **5 = Very positive** | **4** |

**Table A4.18.**
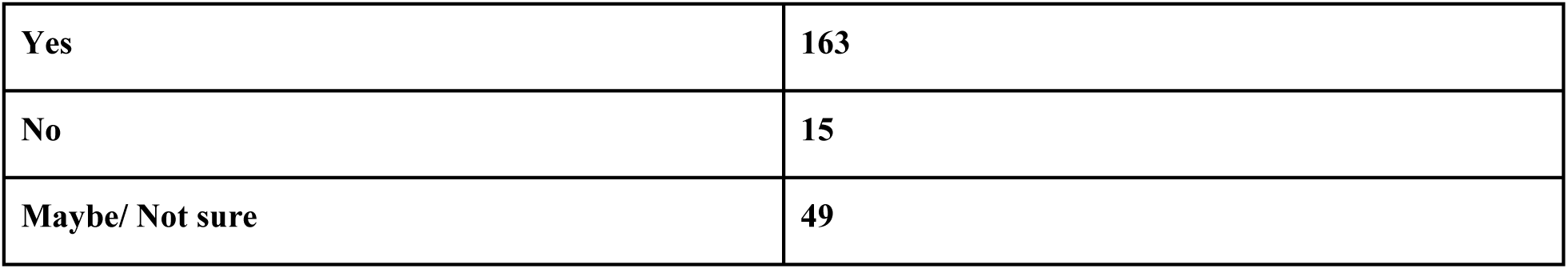
Reponses to question: Are you concerned about the potential impact on training with the expansion of Advanced Practitioner roles?

|  |  |
| --- | --- |
| **Yes** | **163** |
| **No** | **15** |
| **Maybe/ Not sure** | **49** |

Data tables pertaining to non-radiologist doctors in radiology

**Table A4.19.**
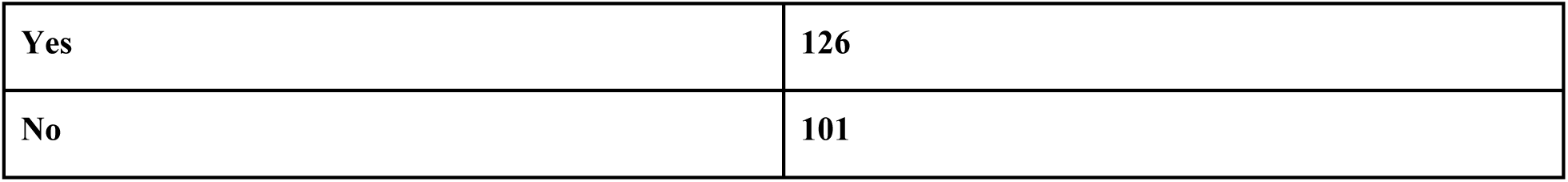
Reponses to question: Have you had experience with non-radiologist doctors reporting imaging studies or image guided procedures?

**Table A4.20.**
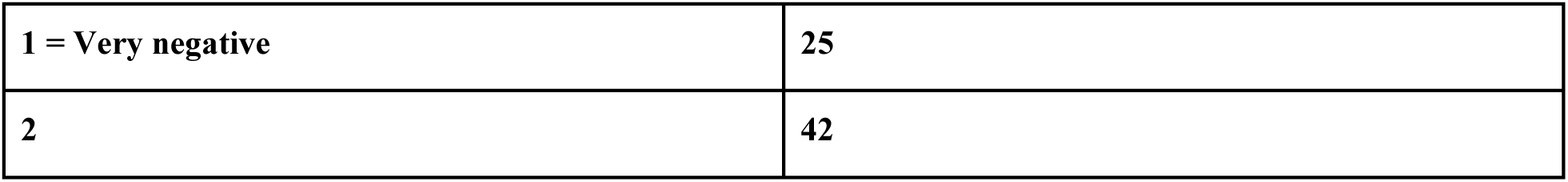
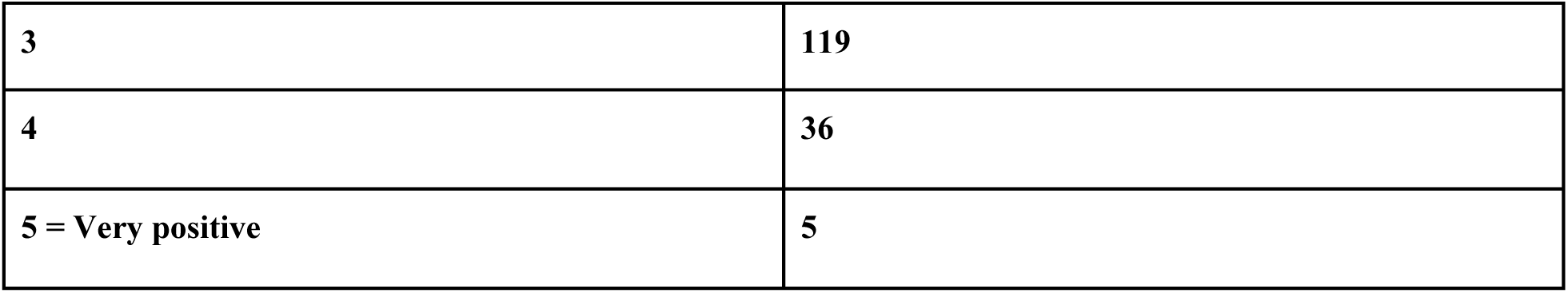
Reponses to question: Non-Radiologist Doctors Reporting imaging or performing Image guided procedures on patient care? (1=very negative; 5=very positive)

|  |  |
| --- | --- |
| **1 = Very negative** | **25** |
| **2** | **42** |
| **3** | **119** |
| **4** | **36** |
| **5 = Very positive** | **5** |

**Table A4.21.**
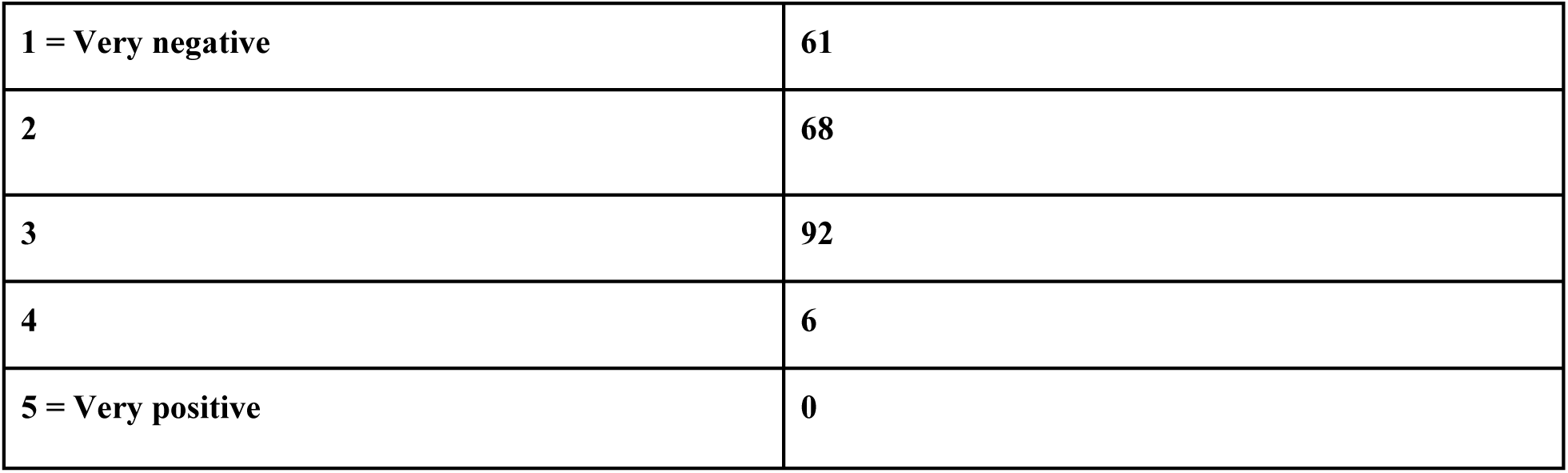
Reponses to question: Non-Radiologist Doctors Reporting imaging or performing Image guided procedures on SpR Training? (1=very negative; 5=very positive)

**Table A4.22.**
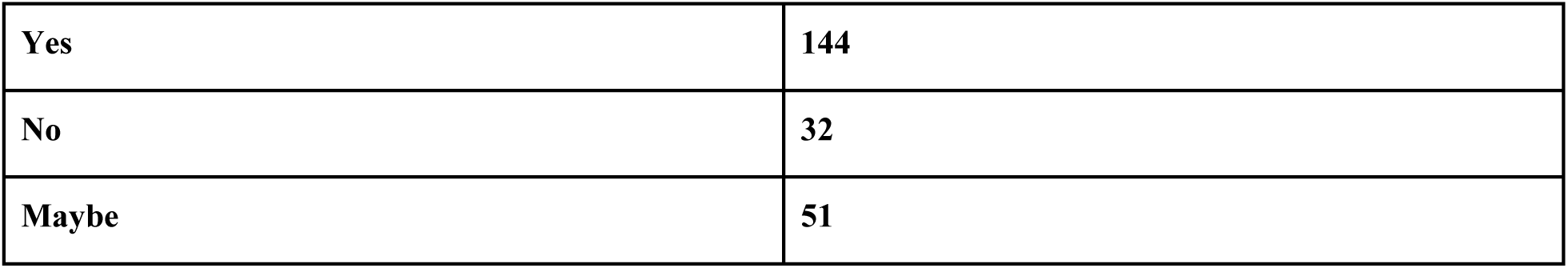
Reponses to question: Are you concerned about the expansion of reporting and intervention to non-radiology doctors?

**Table A4.23.**
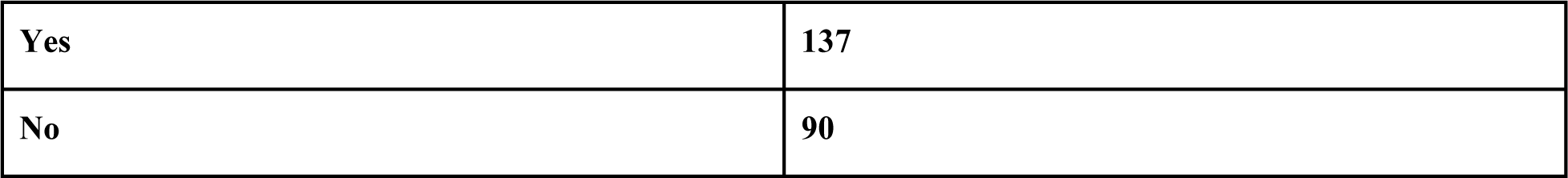
Reponses to question: Have you had experience with using Artificial Intelligence software within your radiology department for image interpretation?

**Table A4.24.**
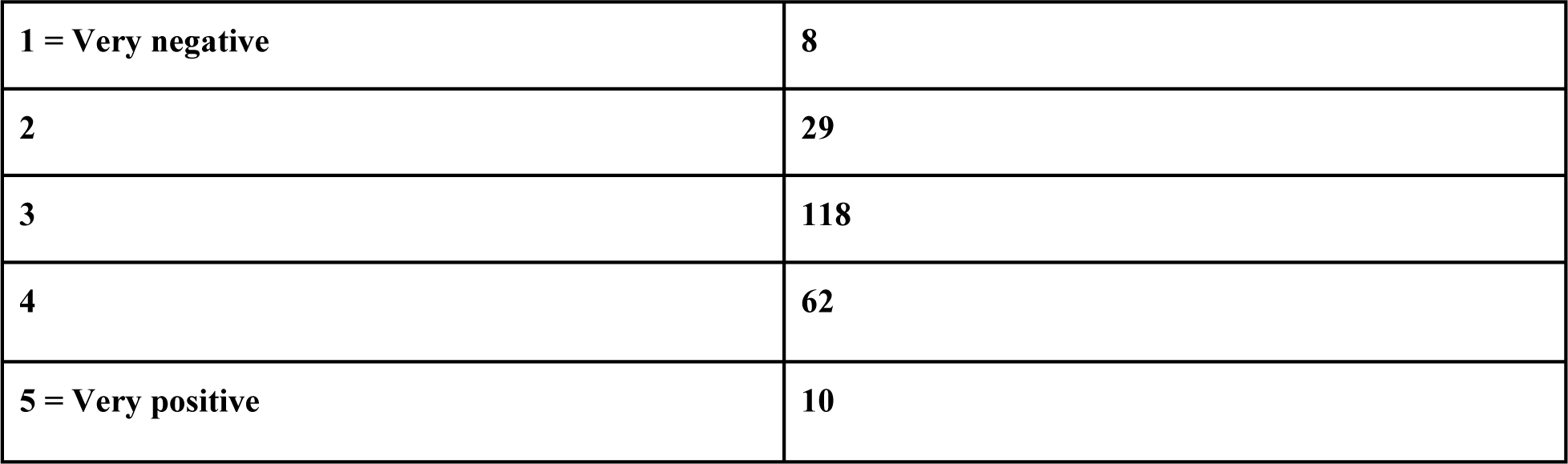
Reponses to question: Effect of AI on Patient care? (1=very negative; 5=very positive)

**Table A4.25.**
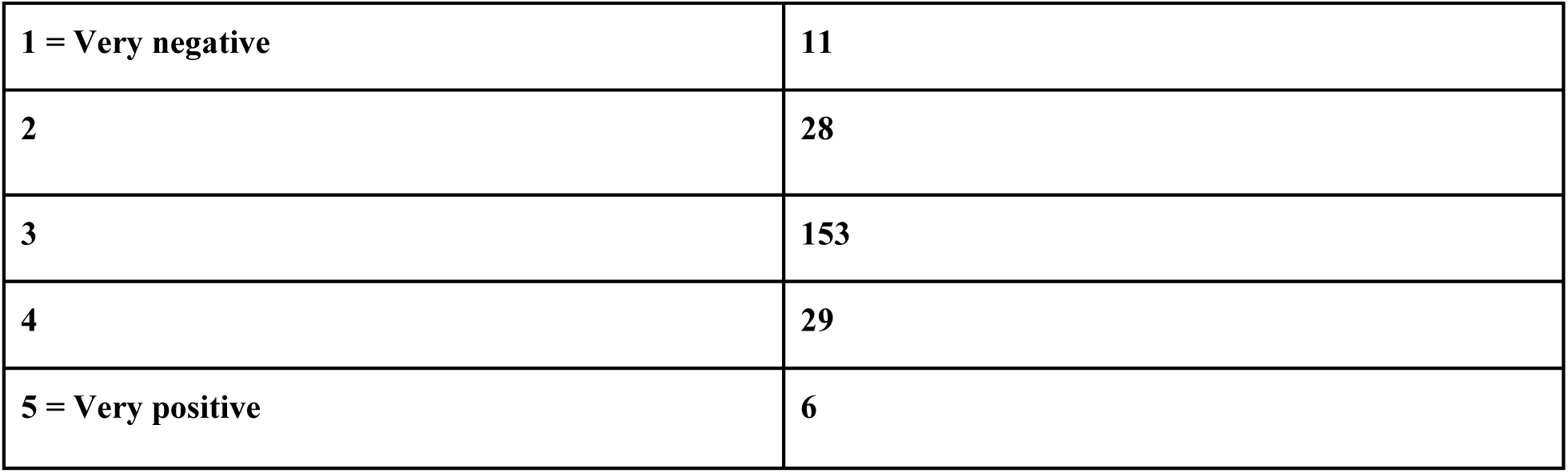
Reponses to question: Impact of AI on SpR Training?(1=very negative; 5=very positive)

## A.5. Future Outlook data tables

**Table A5.1.** Reponses to question: What is your outlook on the future of radiology as a training path/career with expansion of non-radiologist roles and scope?

|  |  |
| --- | --- |
| **1 = Very pessimistic** | **49** |
| **2** | **90** |
| **3** | **47** |
| **4** | **32** |
| **5 = Very positive** | **9** |

**Table A5.2.** Reponses to question: When you are a consultant, to what level would you be comfortable in supervising non-radiologists report or perform image guided procedures?

|  |  |
| --- | --- |
| **1 = Not at all** | **125** |
| **2** | **61** |
| **3** | **27** |
| **4** | **12** |
| **5 = Fully** | **2** |

**Table A5.3.** Reponses to question: As a consultant, to what level would you be happy to train non-radiologists to report or perform image guided procedures?

|  |  |
| --- | --- |
| **1 = Not at all** | **137** |
| **2** | **52** |
| **3** | **22** |
| **4** | **14** |
| **5 = Fully** | **2** |

**Table A5.4.**
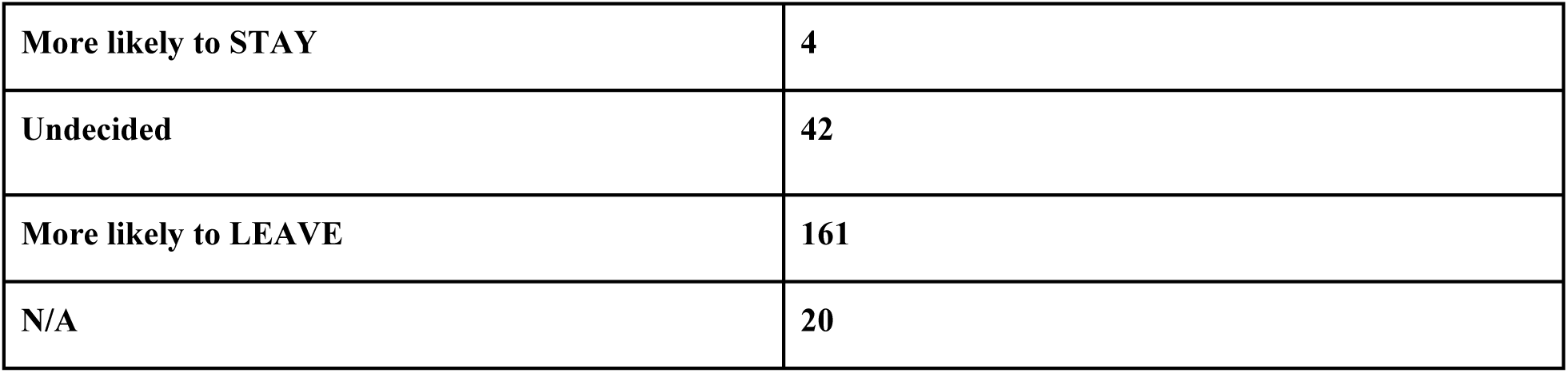
Reponses to question: How will the expansion of the extended practice non-radiology roles impact your decision to stay in the UK after completing training?

